# Risk factors for severe COVID-19 outcomes: a study of immune-mediated inflammatory diseases, immunomodulatory medications, and comorbidities in a large US healthcare system

**DOI:** 10.1101/2023.06.26.23291904

**Authors:** Qi Wei, Prof Philip J. Mease, Michael Chiorean, Lulu Iles-Shih, Wanessa F. Matos, Andrew Baumgartner, Sevda Molani, Yeon Mi Hwang, Basazin Belhu, Alexandra Ralevski, Jennifer Hadlock

**Affiliations:** Institute for Systems Biology, Seattle, WA, USA; Providence St. Joseph Health/ Swedish Medical Center, Rheumatology, Seattle, WA, USA; Digestive Health Institute, Swedish Medical Center, Seattle, WA, USA; University of Washington, Biomedical Informatics and Medical Education, Seattle, WA, USA

**Author notes:** Authors contributed equally (Qi Wei, Prof Philip J. Mease). Preprint submitted to The Lancet. October 31, 2022.

**Keywords:** Autoimmune disease, COVID-19, risk, immunomodulatory medications, multiple chronic conditions

## Abstract

**Background:** COVID-19 outcomes, in the context of immune-mediated inflammatory diseases (IMIDs), are incompletely understood. Reported outcomes vary considerably depending on the patient population studied. It is essential to analyse data for a large population, while considering the effects of the pandemic time period, comorbidities, long term use of immunomodulatory medications (IMMs), and vaccination status.

**Methods:** In this retrospective case-control study, patients of all ages with IMIDs were identified from a large U.S. healthcare system. COVID-19 infections were identified based on SARS-CoV-2 NAAT test results. Controls without IMIDs were selected from the same database. Severe outcomes were hospitalisation, mechanical ventilation (MV), and death. We analysed data from 1 March 2020 to 30 August 2022, looking separately at both pre-Omicron and Omicron predominant periods. Factors including IMID diagnoses, comorbidities, long term use of IMMs, and vaccination and booster status were analysed using multivariable logistic regression (LR) and extreme gradient boosting (XGB).

**Findings:** Out of 2 167 656 patients tested for SARS-CoV-2, there were 290 855 with confirmed COVID-19 infection: 15 397 patients with IMIDs and 275 458 controls (patients without IMIDs). Age and most chronic comorbidities were risk factors for worse outcomes, whereas vaccination and boosters were protective. Patients with IMIDs had higher rates of hospitalisation and mortality compared with controls. However, in multivariable analyses, few IMIDs were rarely risk factors for worse outcomes. Further, asthma, psoriasis and spondyloarthritis were associated with reduced risk. Most IMMs had no significant association, but less frequently used IMM drugs were limited by sample size. XGB outperformed LR, with the AUROCs for models across different time periods and outcomes ranging from 0·77 to 0·92.

**Interpretation:** For patients with IMIDs, as for controls, age and comorbidities were risk factors for worse COVID-19 outcomes, whereas vaccinations were protective. Most IMIDs and immunomodulatory therapies were not associated with more severe outcomes. Interestingly, asthma, psoriasis and spondyloarthritis were associated with less severe COVID-19 outcomes than those expected for the population overall. These results can help inform clinical, policy and research decisions.

**Funding:** Pfizer, Novartis, Janssen, NIH

**MeSH:** D001327, D000086382, D025241, D012306, D000071069

## 1. Introduction

Coronavirus disease (COVID-19) remains a challenge, affecting people worldwide with more than 6·8 million deaths^1^ as of 10 March 2023. Given the variability in the course and outcomes of the disease and its relationship with the immunologic system, understanding COVID-19 outcomes for patients with immune-mediated inflammatory diseases (IMIDs) is essential.

IMIDs are a set of clinically diverse conditions characterized by immune dysregulation, chronic inflammation and potential organ damage. IMIDs include autoimmune diseases, such as rheumatoid arthritis (RA) and multiple sclerosis (MS), as well as inflammatory conditions, including allergic asthma.

Given established and potential COVID-19 risk factors, individuals with IMIDs are of particular interest and complexity for risk analysis.^2,3,4,5^ This population has a higher rate of severe COVID-19 outcomes, but we need to understand why. Potential reasons include immune dysregulation, the use of immunomodulatory drugs (IMMs), and/or associated chronic comorbidities. Moreover, patients with IMIDs have increased rates of comorbidities associated with more severe COVID-19 outcomes, including heart disease and diabetes compared to the general population.^6^ IMMs for IMIDs could theoretically foster viral replication, which could be detrimental in the early stages of disease, but may also reduce the systemic inflammatory response syndrome associated with organ damage, morbidity and mortality.^7^ Also, patients may have multiple IMIDs and/or may be taking multiple IMMs. Allergic asthma has also been shown to be associated with reduced susceptibility to severe COVID-19 outcomes, leading to new insights on the intrinsic factors modulating viral load and spreading mechanisms.^5^

Outcomes must also be considered in the context of changes over the course of the pandemic, including SARS-CoV-2 variants, increasing access to vaccination, and changes in standard of care for COVID-19 treatment.

Previous research suggests that, in addition to advanced age and male sex, specific chronic comorbidities have been associated with higher risk for hospitalisation and death among patients with SARS-CoV-2 and IMIDs, including diabetes mellitus (DM), chronic kidney disease (CKD), chronic obstructive pulmonary disease (COPD), cardiovascular diseases (CVD) and cancer.^8,9,10^ However, risks associated with IMIDs themselves are less well understood. Two large cohort studies in adults compared patients with and without IMIDs and found that mortality was higher in IMID patients.^9,11^ Similarly, some studies have indicated an association between IMMs and high risk for severe COVID-19 outcomes,^12^ whereas others have not.^7,13^

Previous studies analysing IMIDS, IMMs and/or comorbidities^9,11,14,15,16^ did not include Omicron variants or vaccination status or were conducted on smaller cohorts.^17^ A recent study using data from the National COVID Cohort Collaborative (N3C) that spans both pre- and post-Omicron has shown that patients with a prior IMID or prior exposure to IMMs have a higher risk of life-threatening COVID-19 outcomes. However, the study design grouped different IMIDs together, making it difficult to understand the role of individual IMIDs in COVID-19 outcomes.^18^ In this study, we analyse severe outcomes, including hospitalisation, mechanical ventilation (MV) and death, using multivariable models across a large US population to investigate the risk associated with individual IMIDs, classes of IMMs, chronic comorbidities, and vaccination status. The study period was dichotomized so that we could compare the pre-Omicron and Omicron-predominant periods.

## 2. Research in context

### 2.1. Evidence before this study

We searched PubMed, medRxiv, bioRxiv, and Google Scholar for large-population multivariable studies published from 1 January 2020, to 24 April 2023, using the term “COVID-19” with “immune-mediated inflammatory disease”, “autoimmune disease”, “immunomodulatory medication”, “immunosuppressive medication”, and “rheumatic disease”. We also searched for “infection”, “hospitalisation”, “mechanical ventilation”, and “mortality” risk and rates related to COVID-19 in patients receiving immunomodulatory medications. Review was limited to articles published in English. To our knowledge, as of 24 April 2023, there has not yet been a study of risk for severe COVID-19 outcomes that accounts for the interconnected factors of specific IMIDs, classes of immunomodulatory medications, chronic comorbidities, and vaccination status, with consideration of both early pandemic conditions and more recent Omicron variants.

### 2.2. Added value of this study

We developed high accuracy machine learning models on electronic health record (EHR) data for over two million patients who were tested for COVID-19 across seven states in the US, looking separately at two different time frames across the pandemic: before and after the emergence of Omicron. In both time frames, patients with IMIDs had a lower rate of COVID-19 infection but those with infection were more likely to be hospitalised or die when compared with non-IMID controls. Age and most chronic comorbidities were associated with worse outcomes, whereas vaccinations and boosters were associated with better outcomes. Select IMIDs showed some higher risk for severe outcomes, but asthma, psoriasis, and spondyloarthritis were associated with better outcomes than expected for the population overall. Most classes of immunomodulatory medications were not associated with increased risk, but some IMM drugs were limited by sample size and will require further study.

### 2.3. Implications of all the available evidence

Although patients with IMIDs are at higher risk for severe COVID-19, this research suggests that their comorbidities are greater risk factors, rather than the IMIDs themselves, or the use of immunomodulatory drugs. Interestingly, asthma, psoriasis and spondyloarthritis were associated with better outcomes than those expected for the population overall. These results can help inform clinical, policy and research decisions.

## 3. Methods

This retrospective study protocol was reviewed and approved by the Institutional Review Board at Providence Health & Services and affiliates (PHSA) through expedited review (study number: STUDY2021000592). Consent was waived because disclosure of protected health information for the study was determined to involve no more than a minimal risk to the privacy of individuals.

### 3.1. Study setting and participants

In this retrospective case-control study, clinical data was derived from electronic health records (EHRs) from PHSA, an integrated healthcare system which serves patients in 51 hospitals and 1,085 clinics across seven US states: Alaska, California, Montana, Oregon, New Mexico, Texas, and Washington. Patients with IMIDs were identified based on their medical history and all unmatched controls were selected from the same database. Two times frames were analysed: 1 March 2020 to 25 December 2021 (pre-Omicron, when the wildtype, Alpha, Beta, and Delta variants were predominant), and 26 December 2021 to 30 August 2022 (Omicron predominant). The cohort selection flow chart is shown in Figure 1A.

**Figure 1:**
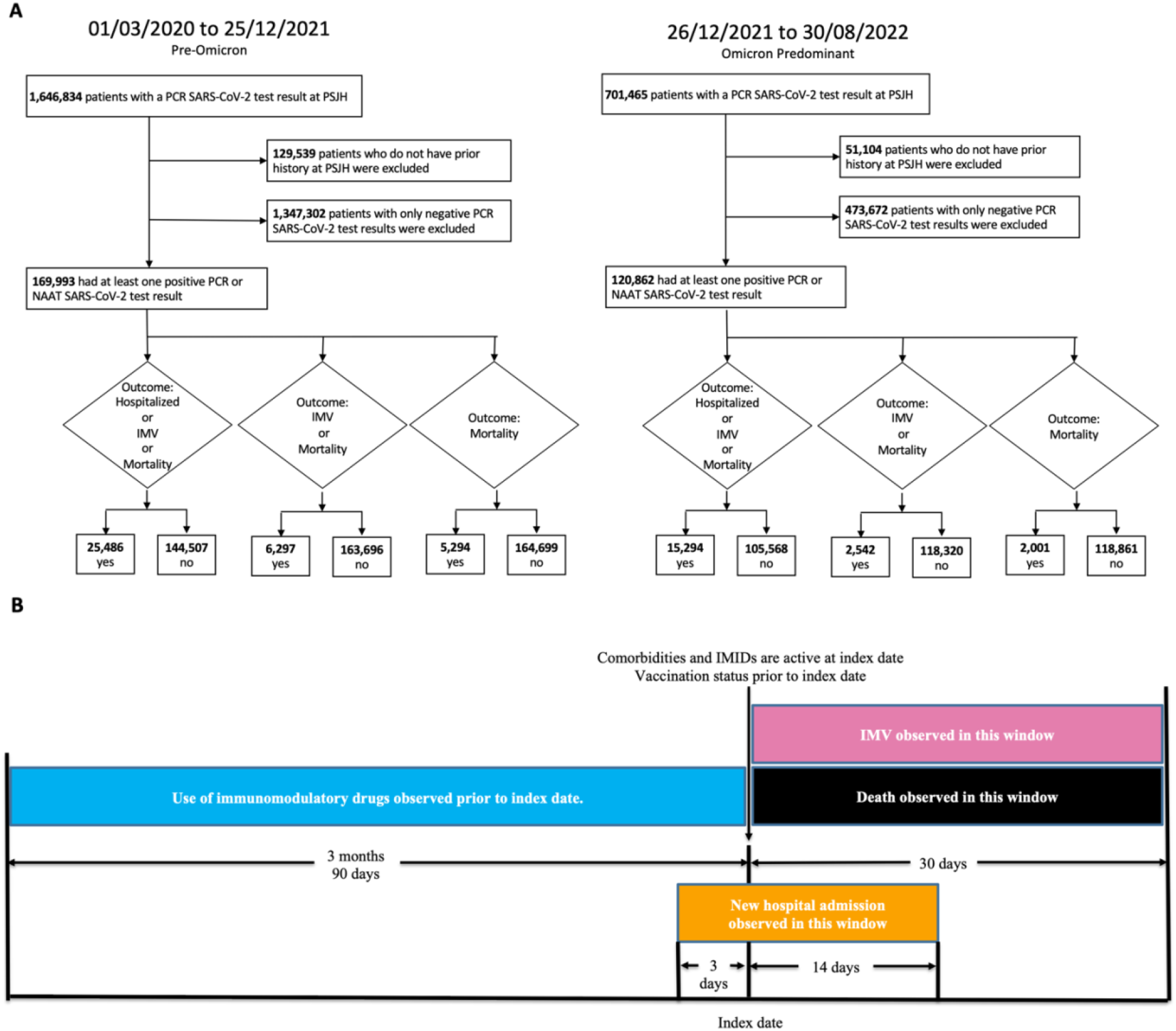
A. Cohort flow diagram for retrospective study of patients at PHSA = Providence Health & Services and affiliates in the U.S. (Alaska, California, Montana, New Mexico, Oregon, Washington, or Texas). B. Timeline for observation of characteristics and outcomes. For patients with COVID-19, the index date was set to their first positive test; For COVID-19 negative patients, the index date was set to their first negative test. Patients with severe COVID-19 outcomes (hospitalisation, MV, and/or death) were identified if they had new hospital admission (orange) within the window of three days prior to 14 days of index date; usage of MV (purple) or death (black) within 30 days of index date. Patients’ use of immunomodulatory drugs was observed for three months leading up to the index date (blue). Patients’ comorbidities and IMIDs’ active status were decided based on the index date. Patients’ vaccination status was decided prior to the index date. For patients with a COVID-19 vaccine from either Moderna or Pfizer, we counted administered doses = 2 as “fully vaccinated” and > 2 as “boosted”. For patients with a COVID-19 vaccine from Janssen, we counted an administered dose = 1 as “fully vaccinated” and > 1 as “boosted”. All vaccination information was obtained from state records and limited to the seven states in our study.

Results were observed for patients who had a valid nucleic acid amplification test (NAAT) for SARS-CoV-2. The index date was considered the date of a COVID-19 test (infection date or first negative test). To be able to determine the effect of the use of IMMs at the time of infection, we selected a subset of patients that had at least one encounter with PHSA prior to the index date. Difference in rates for outcomes was assessed using Fisher’s exact test.

### 3.2. COVID-19 severe outcomes

The following outcomes were evaluated among COVID+ patients: 1) combined endpoint of hospitalisation, MV, and death, 2) combined endpoint of MV and death, and 3) death. See Table 1. Time windows used to define outcomes are shown in Figure 1B.

**Table 1.**
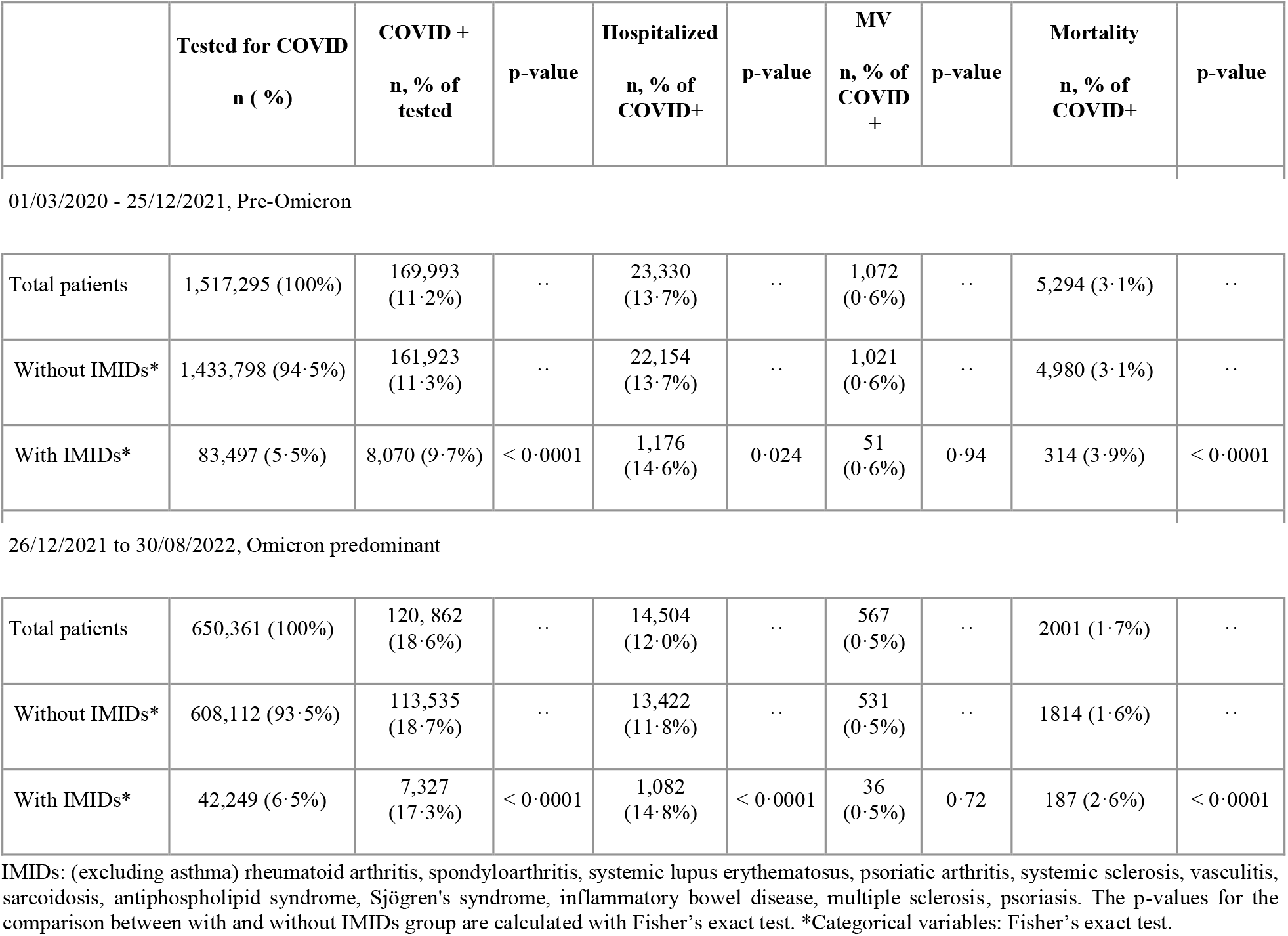
COVID-19 test results, hospitalization, and mortality

|  | **Tested for COVID**<br>**n ( %)** | **COVID +**<br>**n, % of tested** | **p-value** | **Hospitalized**<br>**n, % of COVID+** | **p-value** | **MV**<br>**n, % of COVID +** | **p-value** | **Mortality**<br>**n, % of COVID+** | **p-value** |
| --- | --- | --- | --- | --- | --- | --- | --- | --- | --- |
| 01/03/2020 - 25/12/2021, Pre-Omicron |  |  |  |  |  |  |  |  |  |
| Total patients | 1,517,295 (100%) | 169,993 (11.2%) | .. | 23,330 (13.7%) | .. | 1,072 (0.6%) | .. | 5,294 (3.1%) | .. |
| Without IMIDs* | 1,433,798 (94.5%) | 161,923 (11.3%) | .. | 22,154 (13.7%) | .. | 1,021 (0.6%) | .. | 4,980 (3.1%) | .. |
| With IMIDs* | 83,497 (5.5%) | 8,070 (9.7%) | < 0.0001 | 1,176 (14.6%) | 0.024 | 51 (0.6%) | 0.94 | 314 (3.9%) | < 0.0001 |
| 26/12/2021 to 30/08/2022, Omicron predominant |  |  |  |  |  |  |  |  |  |
| Total patients | 650,361 (100%) | 120,862 (18.6%) | .. | 14,504 (12.0%) | .. | 567 (0.5%) | .. | 2001 (1.7%) | .. |
| Without IMIDs* | 608,112 (93.5%) | 113,535 (18.7%) | .. | 13,422 (11.8%) | .. | 531 (0.5%) | .. | 1814 (1.6%) | .. |
| With IMIDs* | 42,249 (6.5%) | 7,327 (17.3%) | < 0.0001 | 1,082 (14.8%) | < 0.0001 | 36 (0.5%) | 0.72 | 187 (2.6%) | < 0.0001 |

IMIDs: (excluding asthma) rheumatoid arthritis, spondyloarthritis, systemic lupus erythematosus, psoriatic arthritis, systemic sclerosis, vasculitis, sarcoidosis, antiphospholipid syndrome, Sjögren's syndrome, inflammatory bowel disease, multiple sclerosis, psoriasis. The p-values for the comparison between with and without IMIDs group are calculated with Fisher's exact test. \*Categorical variables: Fisher's exact test.

### 3.3. Cohort characteristics

The list of demographics, IMIDs, comorbidities, and IMMs for all tested patients and those with positive results are shown in Table 2. Comorbidities and IMIDs were identified by diagnosis codes, using SNOMED-CT (version obtained on June 27, 2022, Supplementary Table S8). IMMs were identified by RxNorm medication order codes (Supplementary Table S9).

**Table 2.**
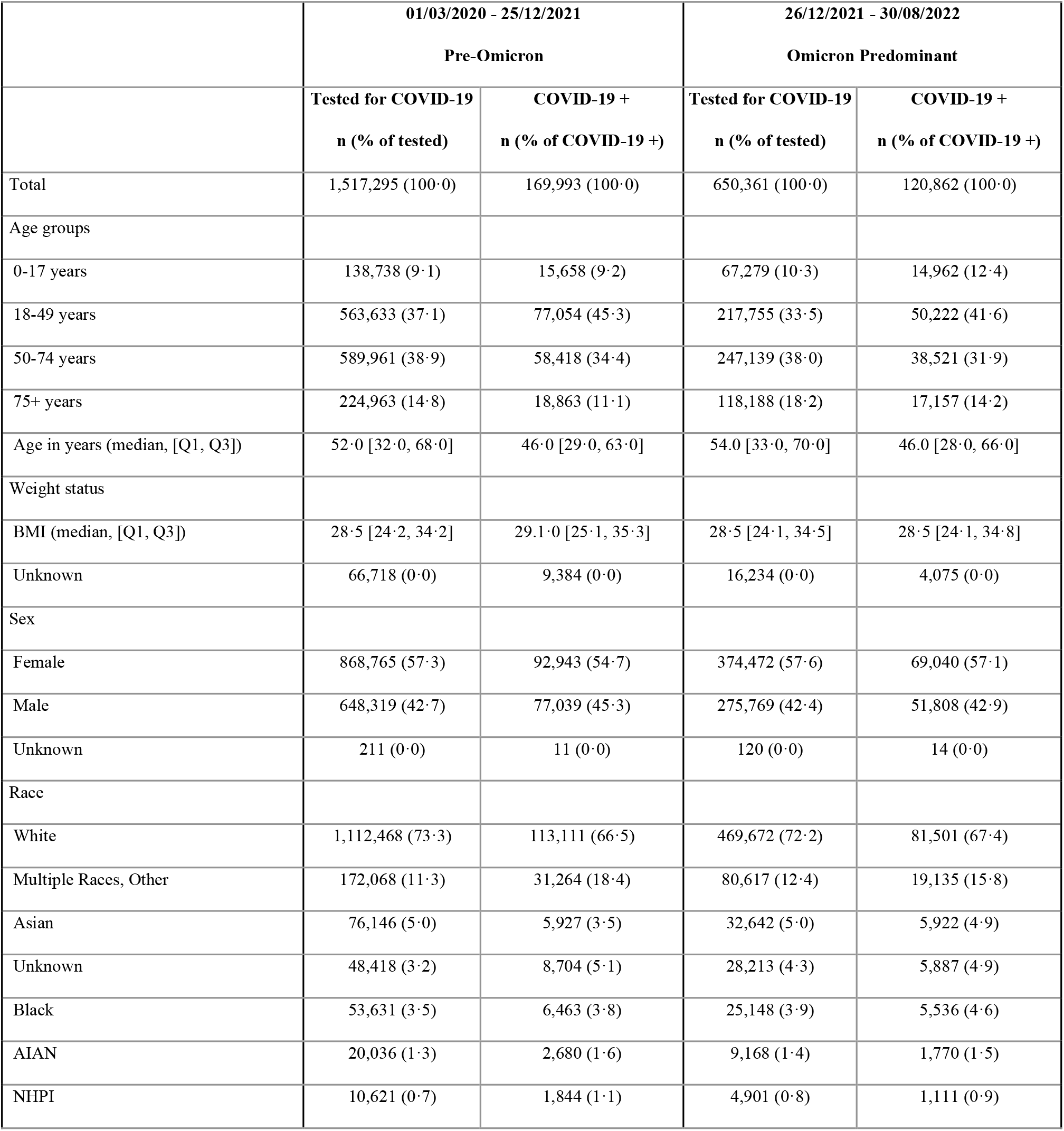
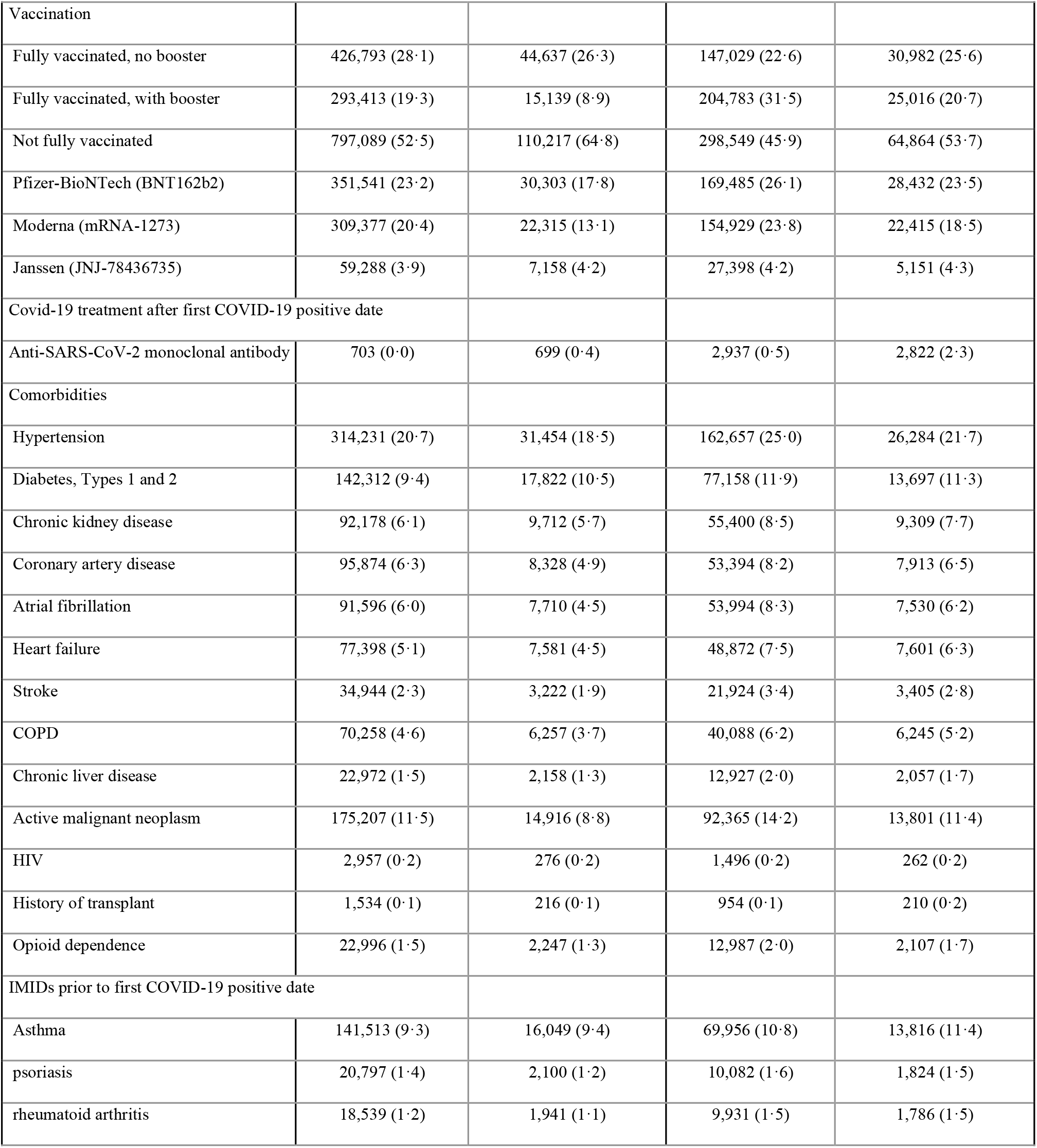
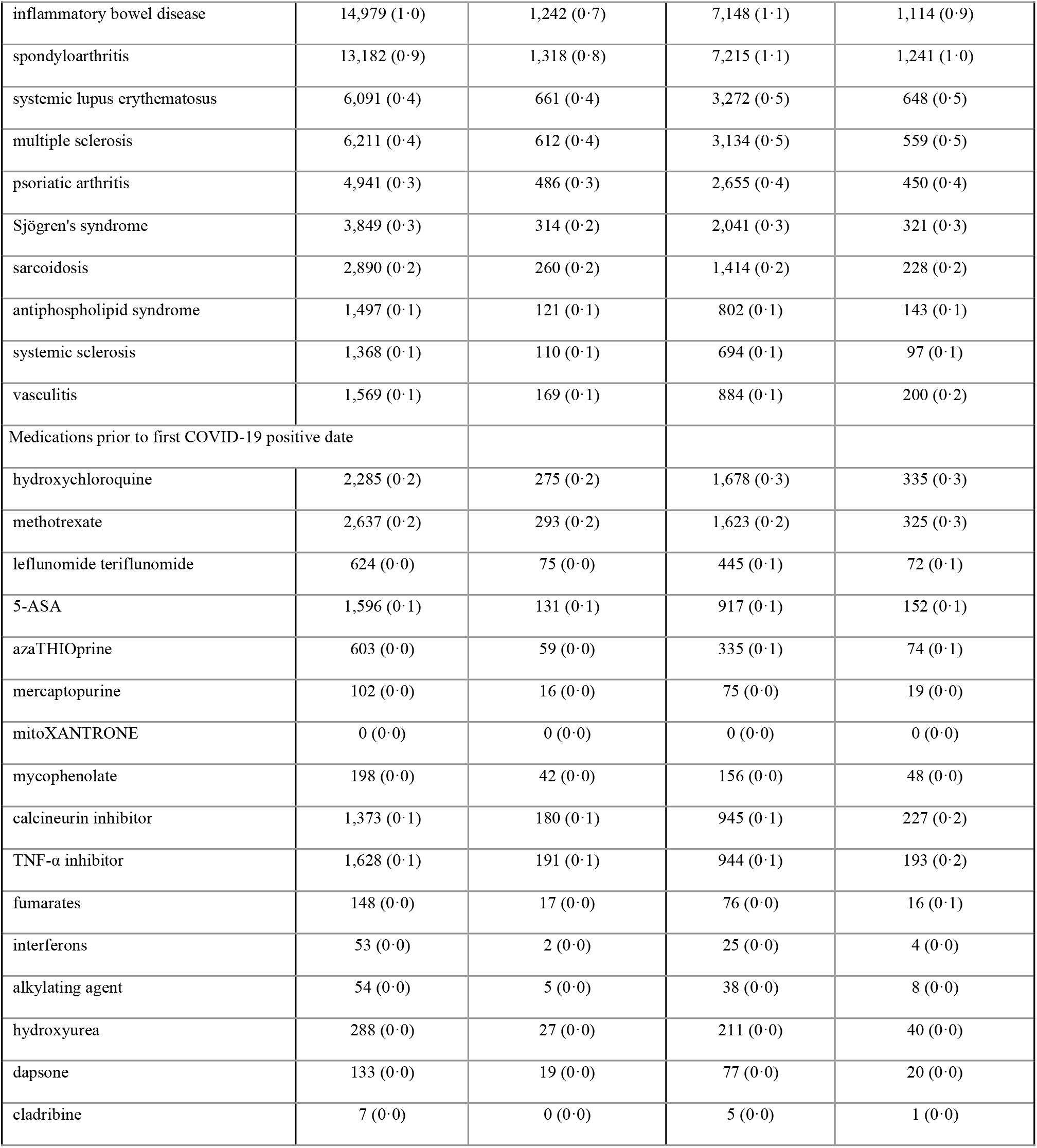
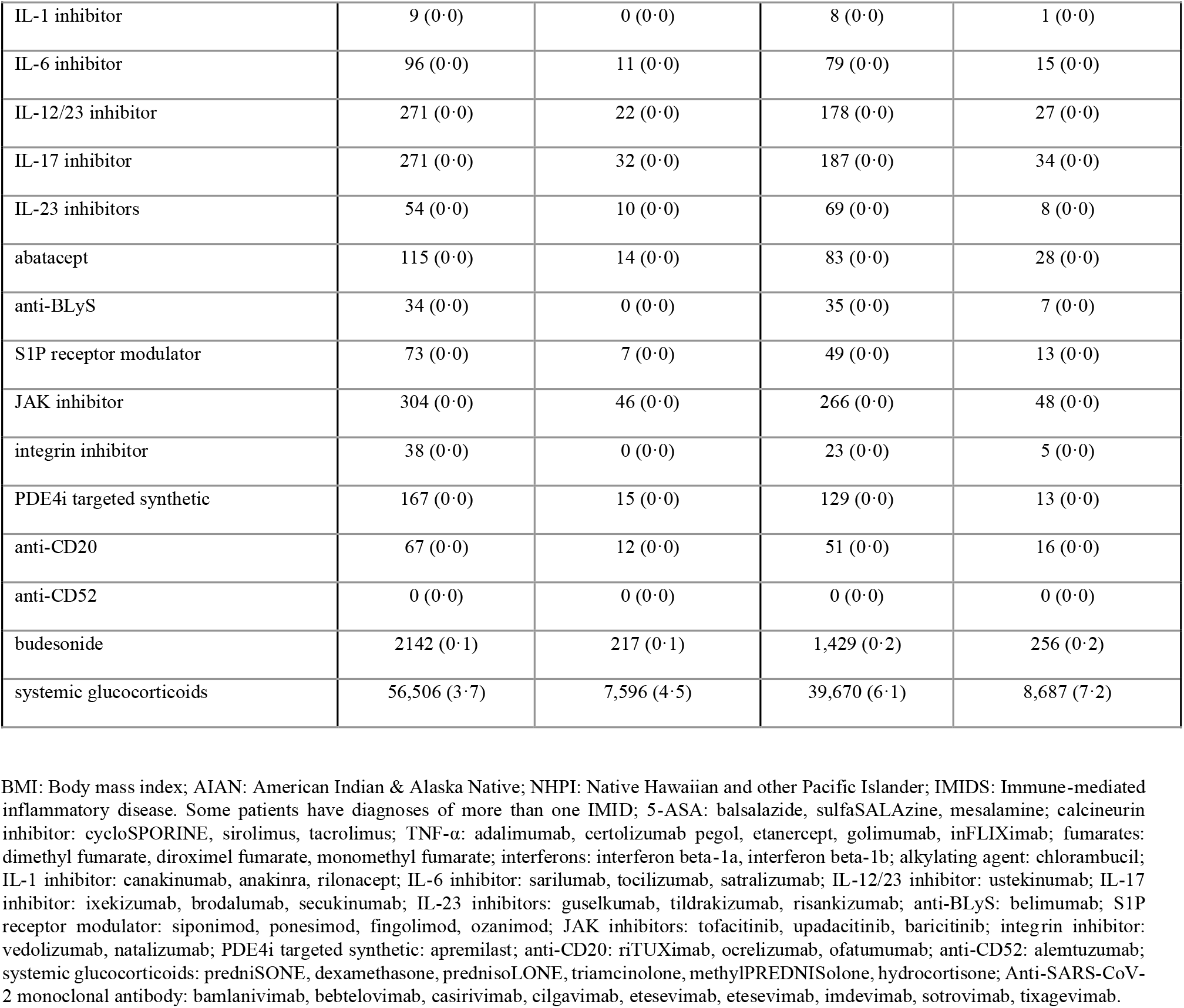
Demographic and clinical characteristics of IMID patients at time of first confirmed SARS-CoV-2 diagnosis

### 3.4. Machine learning models used for classification

To evaluate which variables are most predictive of severe outcomes, we trained supervised learning models on 62 features for two SARS-CoV-2 positive cohorts. Variables included patient demographics, vaccination status, active comorbidities, diagnoses of IMIDs, and use of IMMs within the three months leading up to infection. Time windows used to define variables are shown in Figure 1B. Detailed vaccination definition are shown in caption of Figure 1. Continuous variables, age and body mass index (BMI) were normalized by applying min-max transformation. Missing data were addressed by 1) using the median value to impute missing BMI and 2) assuming absence of active comorbidities, IMIDs, medication usage, and vaccination when those were not reported as active in structured EHR data.

Two alternate analyses were conducted to analyse anti-SARS-CoV-2 neutralizing monoclonal antibodies (NmAbs): one with an additional binary variable for administration of anti-SARS-Cov-2 NmAbs within ten days after COVID-19 test results, and one excluding patients who received anti-SARS-CoV-2 NmAbs. See full results at Supplementary figure S6 and S7. An additional analysis was also conducted with paxlovid. See full results at Supplementary figure S8.

We selected two machine learning methods: 1) traditional logistic regression (LR), for ease of interpretability, and 2) extreme gradient boosted decision tree (XGB), which is an efficient implementation of the regularized gradient boosted decision trees (GBDT) model that can learn non-linear relationships from a high-dimensional datasets and achieve good performance without cost-prohibitive compute requirements. Three additional modelling approaches were assessed to test the assumption that XGB would have the highest performance: adaptive boosting (AdaBoost), K-nearest neighbours (KNN), and support vector machine (SVM). See supplementary figure 1.

Models were generated using the Python package sklearn (version1·1·1) and XGB (version 1·6·1), factors with fewer than ten observations were excluded from the LR model (Table 2), and XGB hyperparameters were tuned with sklearn.model_selection.RandomizedSearchCV (see Supplementary Table S1) using ten-fold cross-validation. This observational study followed the STROBE checklist. See Supplementary table S11.

Models were trained on 90% of the data with an over-sampling method for LR model and an over-weight method (controlled by the scale_pos_weight parameter as shown in the Supplementary Table S1) on minority classes to address class imbalance in training data), with 10% of the data held out for independent performance testing of the final models. Performance was evaluated on the test set for the area under the receiver-operator characteristic curve (AUROC), calculated with the function metrics.roc_auc_score from Python package sklearn (version1·1·1) with parameter average = “weighted”. Models were also evaluated by plotting the log-transformed adjusted odds ratio for each feature.

Feature importance and Shapley additive explanations (SHAP) were applied to understand each variable’s marginal contribution and influence on model prediction using the Python libraries sklearn and SHAP (version 0·37·0), respectively.^19^ Variable independence was assessed using the variance inflation factor (VIF) method. A VIF value equal to one indicates no correlation. A VIF value greater than 5·00 is considered as an indicator that a feature is highly correlated with other features and needs additional feature engineering. Benjamini-Yekutieli multiple hypothesis correction was applied to p-value using Python statsmodel package (version 0·12·2). A p-value less than 0·05 is considered as statistically significant in this study after the hypothesis correction applied.

### 3.5. Computational resources

Data processing and ML was conducted on Azure with Databricks 9·1 LTS (includes Apache Spark 3·1·2). All analysis code was implemented and executed in Python version 3·8·10 except R version 4·1·1, which was used for plotting figures.

## 4. Results

### 4.1. Differences in patient demographics between two time periods in the pandemic

Patient characteristics for all cohorts are shown in Table 2. The majority of people testing positive in both periods were unvaccinated. In the Omicron-predominant period, both patients tested for COVID-19 and those with a positive test results had higher rates for comorbidities and higher rates of vaccination.

### 4.2. Rates of hospitalisation and death in patients with non-asthma IMIDs, compared to those without

In the pre-Omicron period, 11·2% (169 993/ 1 517 295) of people tested positive, and of those, 13·7% were hospitalised, 0·6% were on MV, and 3·1% died. Among patients with IMIDs who underwent testing, 9·7% (8 070 / 83 497) were positive for COVID-19. Of these, 14·6% were hospitalised, 0·6% were on MV, and 3·9% died, higher rates than the overall population. In the Omicron-predominant period, the overall rate of positive tests increased to 18·6% (120 862 / 650 361) but the rate for hospitalisation (12·0%), MV (0·5%) and death (1·7%) decreased. Again, in this period, 17·3% (7 327 / 42 249) of patients with IMIDs who tested positive had increased rates of hospitalisation (14·8% vs. 11·8%) and death (2·6% vs. 1·6%) compared to non-IMID patients. All previously mentioned differences between patients with and without IMIDs were statistically significant.

### 4.3. Multivariable logistic regression (LR) analysis

The VIF method showed sufficiently low correlation of all variables. Age during the Omicron-predominant period had the highest value at 3·58. In both periods, age was a significant risk factor for worse outcomes, as were most comorbidities: heart failure, kidney disease, liver disease, COPD, malignancy, atrial fibrillation (AFib), coronary artery disease, stroke, and diabetes. Vaccination and booster status were significantly associated with better outcomes. Psoriasis showed reduced risk for all three outcomes in both time periods, with adjusted odds ratio (AOR) ranging from 0·52 (95% confidence interval (CI) 0·48-0·56) to 0·80 (0·74-0·87). Asthma showed reduced risk for all three outcomes in both periods, with AOR ranging from 0·33 (95% CI 0·32-0.34) to 0·49 (0·48-0·51). Spondyloarthritis was associated with reduced risk in the pre-Omicron period, with AOR ranging from 0·49 (95% CI 0·44-0·54) to 0·67 (0·61-0·74). See full results in Figure 2 and Supplementary Tables S2-S7.

**Figure 2:**
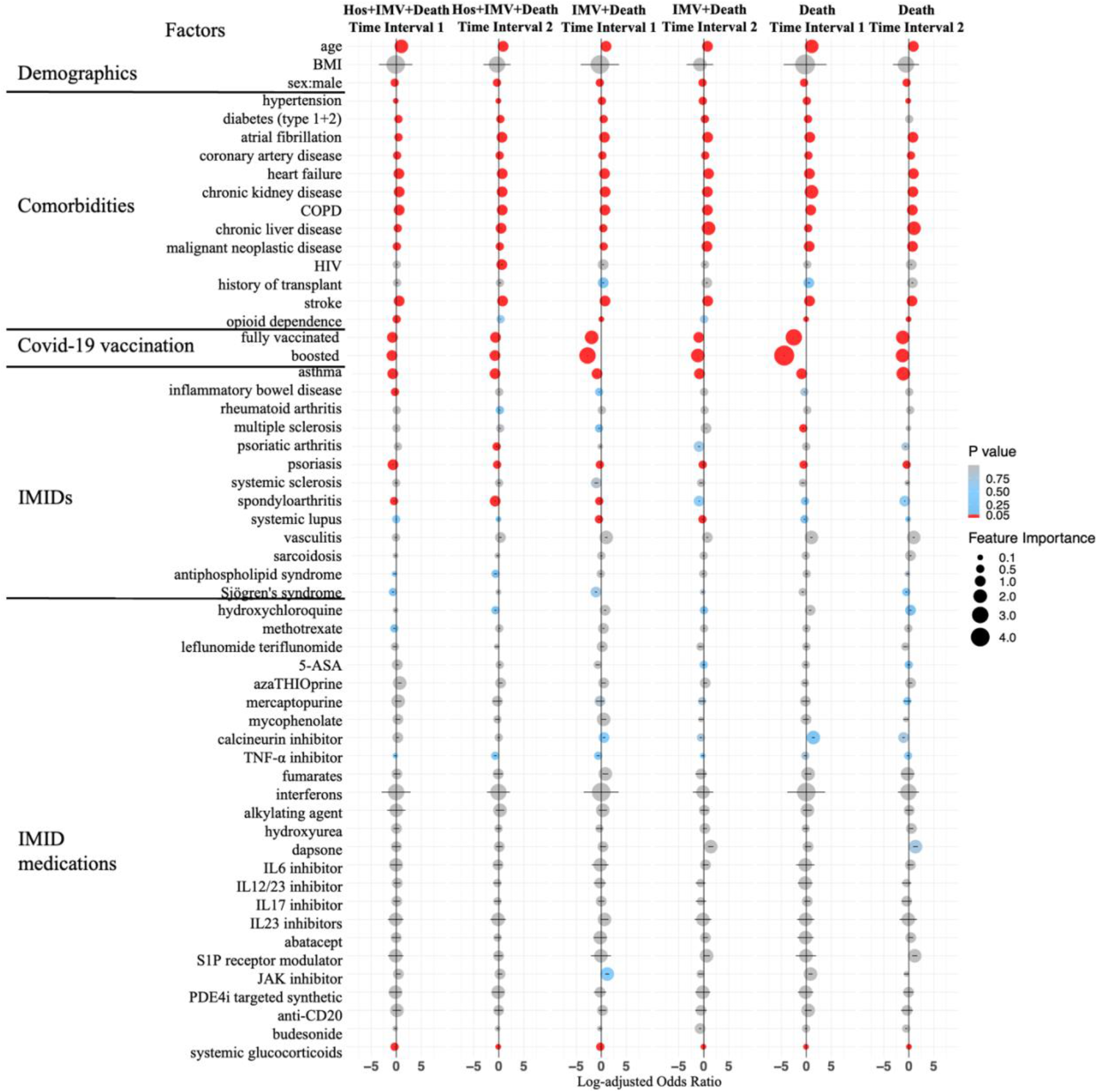
Log adjusted odds ratio. Selected factors for hospitalisation or MV or death, MV or death, and death across Time Interval 1 (01/03/2020 to 25/12/2021) and Time Interval 2 (26/12/2021 to 30/08/2022) in COVID-19 positive patients, using multivariable logistic regression (LR). The colour of dots represent p-values calculated using raw data. Position and size of dots represents log-adjusted odds ratio and feature importance from over-sampling. The error bar represents the 95% confidence interval. Factors with fewer than ten observations were excluded.

### 4.4. XGB and SHAP analysis

XGB had an average of 7·5% better classification performance on the reserved test set compared with LR, across all three outcomes and both time periods (See Supplementary Figure S1). The majority of results, age, most cormorbidities, vaccination, and boosters from SHAP analysis on the XGB model showed similar associations and relative feature importance as seen in LR. (Figures 3-4, and Supplementary figures S3-5). As with LR model, results predicted better outcomes for patients with asthma, spondyloarthritis and psoriasis. XGB and SHAP also illuminated additional results: opioid dependence was predictive of all severe outcomes in both periods, while RA, MS and vasculitis were predictive of some outcomes in the pre-Omicron period, and all three severe outcomes in the Omicron-predominant period. Long term systemic glucocorticoid use had mixed results for hospitalisation but was predictive for MV in the Omicron-predominant period and for death in both time periods.

**Figure 3:**
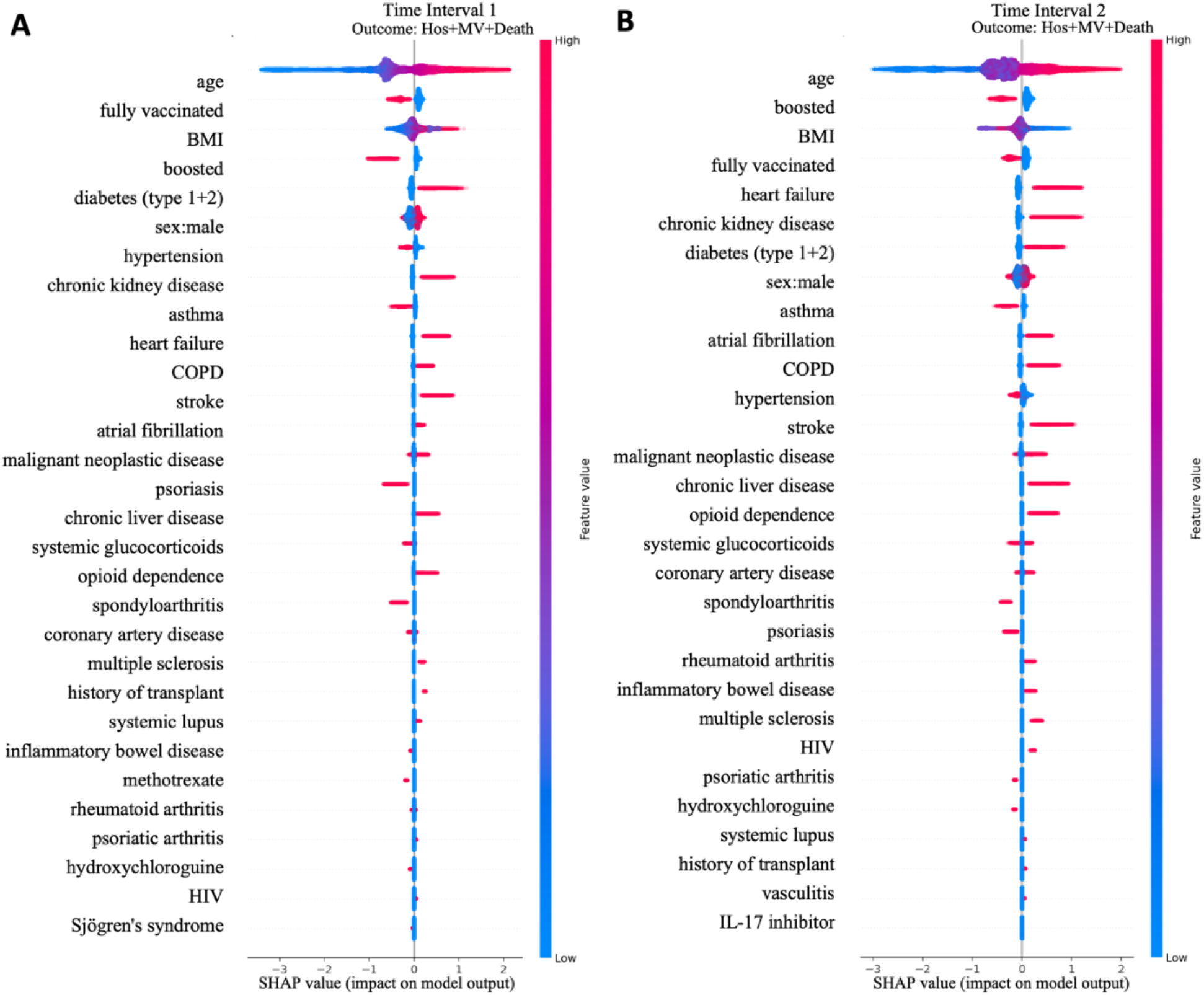
Feature importance for top 30 features of all-age model of hospitalisation within the window of three days prior to 14 days of their first COVID-19 incidence or usage of hospitalisation or death within 30 days of their first COVID-19 incidence across Time Interval 1 (01/03/2020 to 25/12/2021) and Time Interval 2 (26/12/2021 to 30/08/2022). Gradient boosting decision tree feature importance and influence of higher and lower values of the risk factors on the all-age group population outcome. In both A and B plots, SHAP values < 0 are associated with reduced risk for hospitalization or death, and SHAP values > 0 are associated with increased risk. Red dots represent patients with higher values for a variable, and blue dots are patients with lower values. Nominal classes are binary [0, 1]. For sex, male is 1 (red).

## 5. Discussion

In this large retrospective case-control study, similar to other studies, we demonstrated that patients with IMIDs had lower infection rates but higher rates of severe COVID-19 outcomes. Having a specific IMID was less predictive of severe outcomes than age, while vaccination and booster status were protective. RA, vasculitis, and MS had some predictive value for all three severe outcomes during the Omicron-predominant period.

Asthma was associated with reduced risk for all three outcomes, relative to the population overall, supporting previous research about its protective effect.^5^ Interestingly, psoriasis also showed reduced risk for all three outcomes in both time periods, and spondyloarthritis was associated with reduced risk in the pre-Omicron period. The latter adds new evidence to support the spondyloarthritis finding reported earlier by Raiker et al.^20^ In contrast, Rosenbaum et al. reported a small increased risk of developing COVID-19 for patients with spondyloarthritis in the pre-Omicron period but mentioned the risk was not consistently demonstrated.^21^ While this might reflect unmeasured variables, such as behaviours taken to avoid risk of infection, the results merit further investigation into potential disease-related protective factors, such as HLA-B27.^22^

The most important factors for the combined endpoint of hospitalisation, MV or death were age, diabetes, CKD, COPD, stroke, AFib, liver disease, and opioid dependence. The strongest risk factors for mortality were age, pre-existent heart failure, CKD, AFib, COPD, stroke, and liver disease. High BMI has been associated with worse outcomes, but SHAP results on gradient boosting reveal mixed results (Figs. 3,4). This could suggest risk from both very low and very high BMI, or may reflect that the predictive importance of BMI differs between younger and older patients.^23^ Our results confirm previously reported studies on the risks of comorbidities and benefits of vaccination with COVID-19 in the pre-Omicron period.^10,24,25^ We have also shown that these associations continue with Omicron, and that boosters are also predictive of better outcomes.

**Figure 4:**
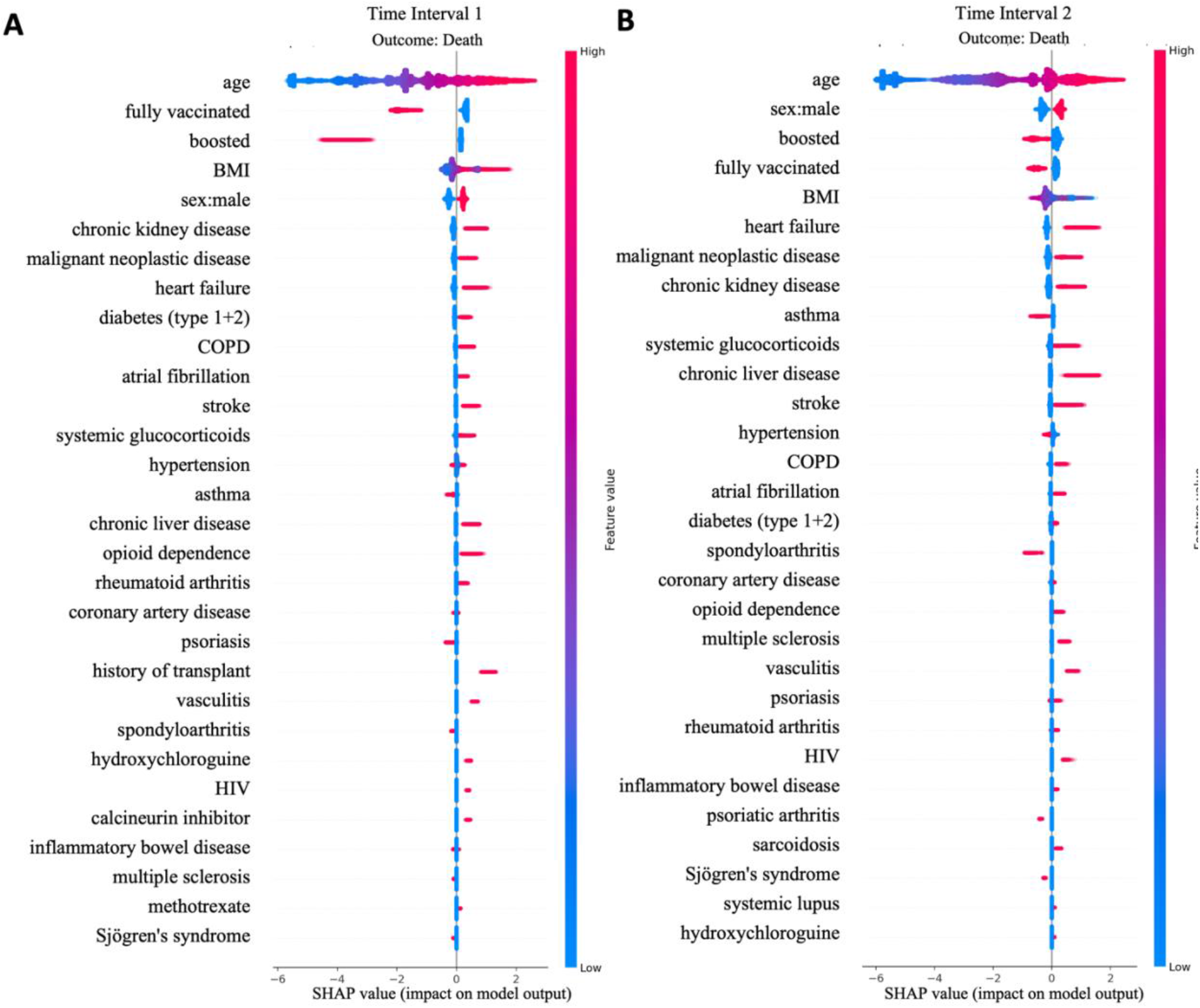
Feature importance for top 30 features of all-age model of death within 30 days of their first COVID-19 incidence across Time Interval 1 (01/03/2020 to 25/12/2021) and Time Interval 2 (26/12/2021 to 30/08/2022). Gradient boosting decision tree feature importance and influence of higher and lower values of the risk factors on the all-age group population outcome. The left side of each plot is associated with reduced risk of death, and the right side of each plot is associated with increased risk of death. Red dots represent patients with higher values for a variable, and blue dots are patients with lower values. Nominal classes are binary [0, 1]. For sex, male is 1 (red).

A UK nationwide cohort study in the OpenSAFELY platform showed COVID-19 autoimmune diseases such as RA, systemic lupus erythematosus (SLE), and psoriasis in the pre-Omicron period^26^ had a higher association of death. We saw some increased risk in some analyses with RA. However, we found better outcomes for patients with psoriasis in both periods. Of note, Piaserico et al. evaluated the quality of earlier studies related to outcomes of COVID-19 in patients with psoriatic arthritis (PsA) and psoriasis treated with IMMs and found risk of bias.^27^

Overall, our results support previously noted associations between specific IMIDs and outcomes from earlier in the pandemic,^6,25,28^ add new insights into areas where there has been uncertainty, provide new results for the Omicron-predominant period, and suggest priorities for deeper investigation into the increased risks observed with RA, MS, and the reduced risks (and potential protective benefits) associated with psoriasis and spondyloarthritis.

Certain IMMs were previously reported to be associated with worse outcomes.^24,25,29,30^ Long term glucocorticoid use only showed an AOR of 1.02, CI:[0.98,1.06] for death during the Omicron-predominant period in LR results but showed predictive value for higher mortality in XGB results for both periods. This supports earlier reports that long-term systemic glucocorticoid use may be implicated in adverse outcomes for some patients with IMIDs in the pre-Omicron period.^10,14,30,31^However, further research is needed on IMID severity at the time of infection, as disease flares may be a confounding factor, affecting both steroid treatment and COVID-19 outcomes. Unfortunately, this information is difficult to extract at scale from structured EHR data. Long term use of other IMMs was not clearly predictive of worse outcomes. However, analysis of less frequently used IMM drugs was limited by sample size and will require further study. For example the number of COVID-19 positive patients on anti-CD20 was very low (only 12 cases in the pre-Omicron period and 16 in the Omicron predominant period).

The results of our study provide support for the conclusion that, for patients with IMIDs, comorbidities and vaccination status are more important predictive factors of COVID-19 outcomes than long-term use of IMMs. However, care should be taken in interpreting population-wide results. Correcting for false discovery rate with multiple hypothesis testing reduces the risk of reporting a spurious signal but increases the risk of overlooking factors that may be important for a subset of patients. For example, other large population studies have noted increased risk of severe COVID-19 outcomes with long term used of rituximab, an anti-CD20 IMM.^25,29,32^

The strengths of our study include having a large population sample size, multivariable analysis with a focus on immune-related risk factors, and application of two complementary modelling approaches. We also investigated the full pandemic time period, comparing the early phase with results after the emergence of Omicron variants to provide insight into whether risk factors have changed since the emergence of Omicron.

Limitations of this study include: a lack of information on 1) IMID disease severity and activity, 2) severity of non-IMID comorbidities, 3) people who had SARS-CoV-2 infection but were not tested at PHSA, 4) several aspects of SARS-CoV-2 immunity, including infection-induced immunity, diminishing effects from immunization over time, and differences in immunity to different variants, 5) sequencing data on actual variants, 6) intensity or dosage of IMMs, 7) rate of IMID underdiagnoses or billing code errors 8) articles published in language other than English. There was also no distinction between medication dosing and or differences in total timing and duration of medications during the three months leading up to the SARS-CoV-2 infection, which is of particular importance for systemic glucocorticoids. Differences in standard of care treatment for COVID-19 were partially accounted for by having two time periods, but the analysis did not include COVID-19 treatment medications at the per-patient level (other than the additional analysis for anti-SARS-CoV-2 NmAbs), and patients with IMIDs may have been treated differently given clinician concern for increased risk. Also, particularly in the Omicron-predominant period, at-home rapid tests became available, leading to a lower actual rate of severe outcomes compared to those reported here.

Although variance inflation factors were considered sufficiently low (below 5·00), age was 3·58 in the Omicron-predominant period, suggesting opportunities for future research into age-stratified models.^27^ Furthermore, we may also have missed other potentially confounding factors such as behavioural choices for patients with immune-related conditions and medications, other socioeconomic exposures, and delays in access to care. In addition, some sample bias may have been introduced when selecting patients who had previously received care at PHSA. This was needed to determine long-term use of IMMs but may have excluded patients, including those who had barriers to accessing care and only sought COVID-19 treatment if they were extremely ill.^33^

## 6. Conclusion

In this paper, we analysed 2 167 656 patients tested for SARS-CoV-2 infection from a large US health care system database regarding risk of COVID-19 infection. For patients with a positive test result, we developed predictive models for severe COVID-19 outcomes (hospitalisation, IMV and death) on 15 397 patients with IMIDs and 275 458 unmatched controls. We examined two separate time periods of the pandemic: pre-Omicron and Omicron-predominant. Patients with IMIDs had higher rates of hospitalisation and mortality than those without. Using multivariable LR and XGB models, we demonstrated that age and most chronic comorbidities were risk factors for severe outcomes, whereas vaccination and boosters were associated with reduced risk. However, apart from RA and MS, specific IMIDs themselves did not show association with worse outcomes. Interestingly, spondyloarthritis and psoriasis were associated with better outcomes, which suggests that, like asthma,^8^ these IMIDs might point to new insights on protective mechanisms against infectious disease.

IMMs were not significantly associated with worse outcomes. Certain classes of immunomodulatory medications showed some predictive value, but due to lack of statistical significance after false discovery rate correction or inconsistency across analytic methods and time periods, these observations need further investigation, and some IMMs may have clinically significant benefit or harm for a subset of patients. More generally, care should be taken in interpretation of all results. Given unmeasured potential confounders, such as those noted in limitations, risk factors are not necessarily causal. However, the results from these more comprehensive multivariate models can help inform clinical, policy and research decisions for patients with IMIDs during the ongoing pandemic.

## Data Availability

All clinical logic has been shared. Results have been aggregated and reported within this paper to the extent possible while maintaining privacy from personal health information as required by law. All data are archived within Providence St Joseph Health systems in a HIPAA- secure audited compute environment to facilitate verification of study conclusions.

https://github.com/Hadlock-Lab/Risk_factors_for_severe_COVID19_outcomes_a_study_of_IMIDs_medications_comorbidities

## Contributors

QW, PJM, MC, LIS, and JJH conceptualised the study. JJH and PJM supervised the study implementation. Funding, administrative support and material support was provided by JJH. QW conducted data cleaning, transformation, statistical analysis and machine learning. AB, YMH, SM, and BB were involved in data interpretation and preparation. QW, WFM, PJM, and JJH prepared the manuscript with critical revision of the manuscript for important intellectual content. QW, PJM, MC, LIS, WFM AB, YMH, SM, BB, and JJH had full access to the data in the study, reviewed and approved the final version of the manuscript and had final responsibility for the decision to submit for publication.

## Data sharing

All clinical logic has been shared. Results have been aggregated and reported within this paper to the extent possible while maintaining privacy from personal health information as required by law. All data are archived within Providence St Joseph Health systems in a HIPAA-secure audited compute environment to facilitate verification of study conclusions.

## Code Availability

The code for extracting, cleaning, and analysing the data in this paper is available on GitHub (https://github.com/Hadlock-Lab/Risk_factors_for_severe_COVID19_outcomes_a_study_of_IMIDs_medications_comorbidities).

## Acknowledgement

This work was funded in part by grants from Pfizer Inc., Novartis Pharmaceutical Corporation, and Janssen Scientific Affairs, LLC. Support was also provided in part by the National Center for Advancing Translational Sciences, National Institutes of Health, through the Biomedical Data Translator program, award #3OT2TR00344301S1. Any opinions expressed in this document are those of the Translator community at large and do not necessarily reflect the views of NCATS, individual Translator team members, or affiliated organizations and institutions.

We are grateful to Providence St Joseph Health for sharing their data engineering expertise and computational resources. We are also grateful for the review of tables and figures done by Evan Yip and cloud computing support by Andrey Dubovoy. We would also like to acknowledge SNOMED International for developing and maintaining SNOMED-CT©.

