## Supplementary Material for "Risk factors for severe COVID-19 outcomes: a study of immune-mediated inflammatory diseases, immunomodulatory medications, and comorbidities in a large US healthcare system"

### Supplementary Figures

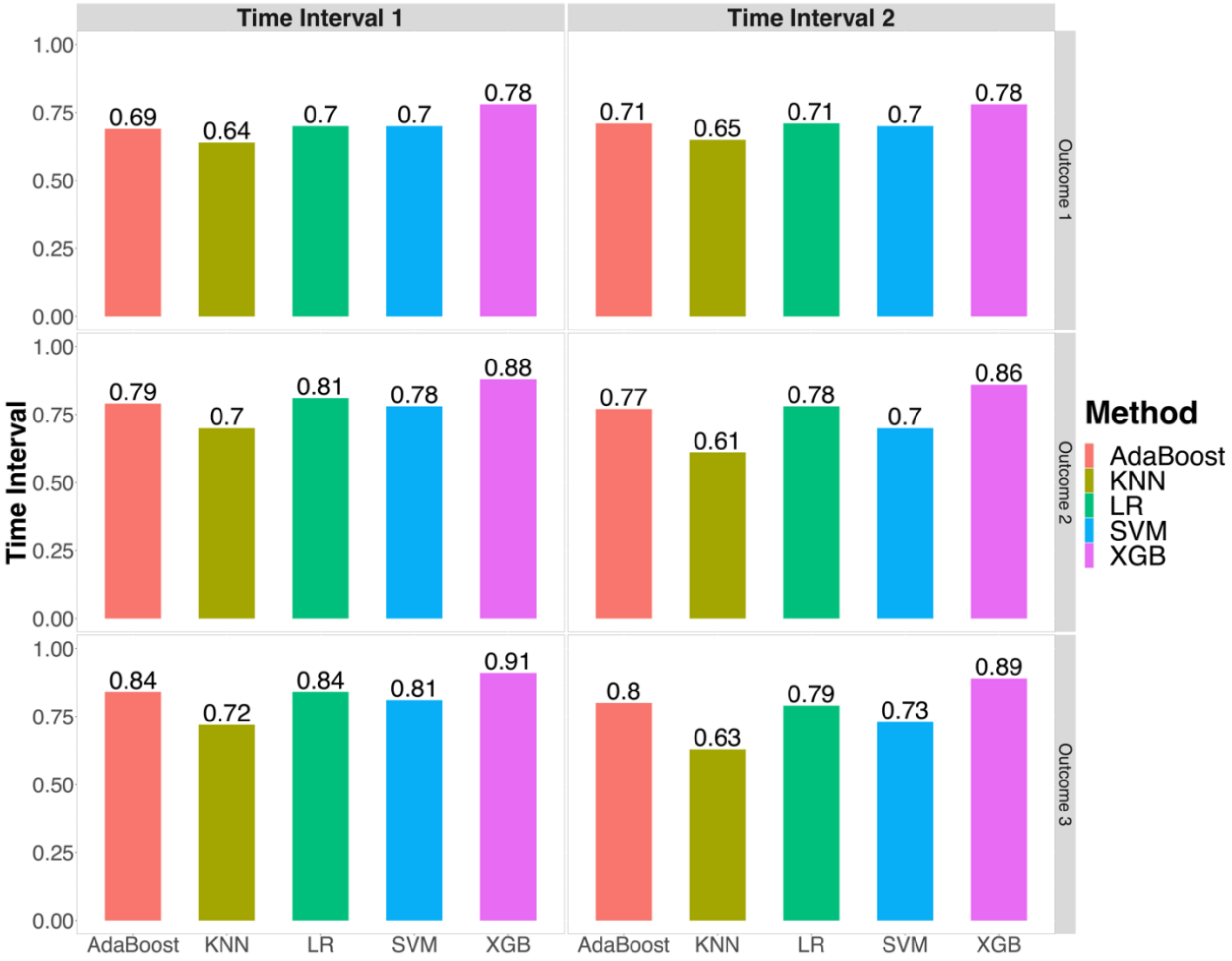

**Supplementary Figure S1. Machine learning models performance on the 10% reserve test set.** Performance for machine learning models for severe outcomes (1. hospitalisation or MV or death, 2. MV or death, and 3. death) across Time Interval 1 (01/03/2020 to 25/12/2021) and Time Interval 2 (26/12/2021 to 30/08/2022)· AUROC = area under the receiver-operator characteristic curve, LR = logistic regression, XGB = gradient boosting, AdaBoost = Adaptive Boosting, KNN = K-nearest neighbours, and SVM = Support Vector Machine. Details of each models' hyperparameters' name and values can be found in Supplementary Table S1 and S12.

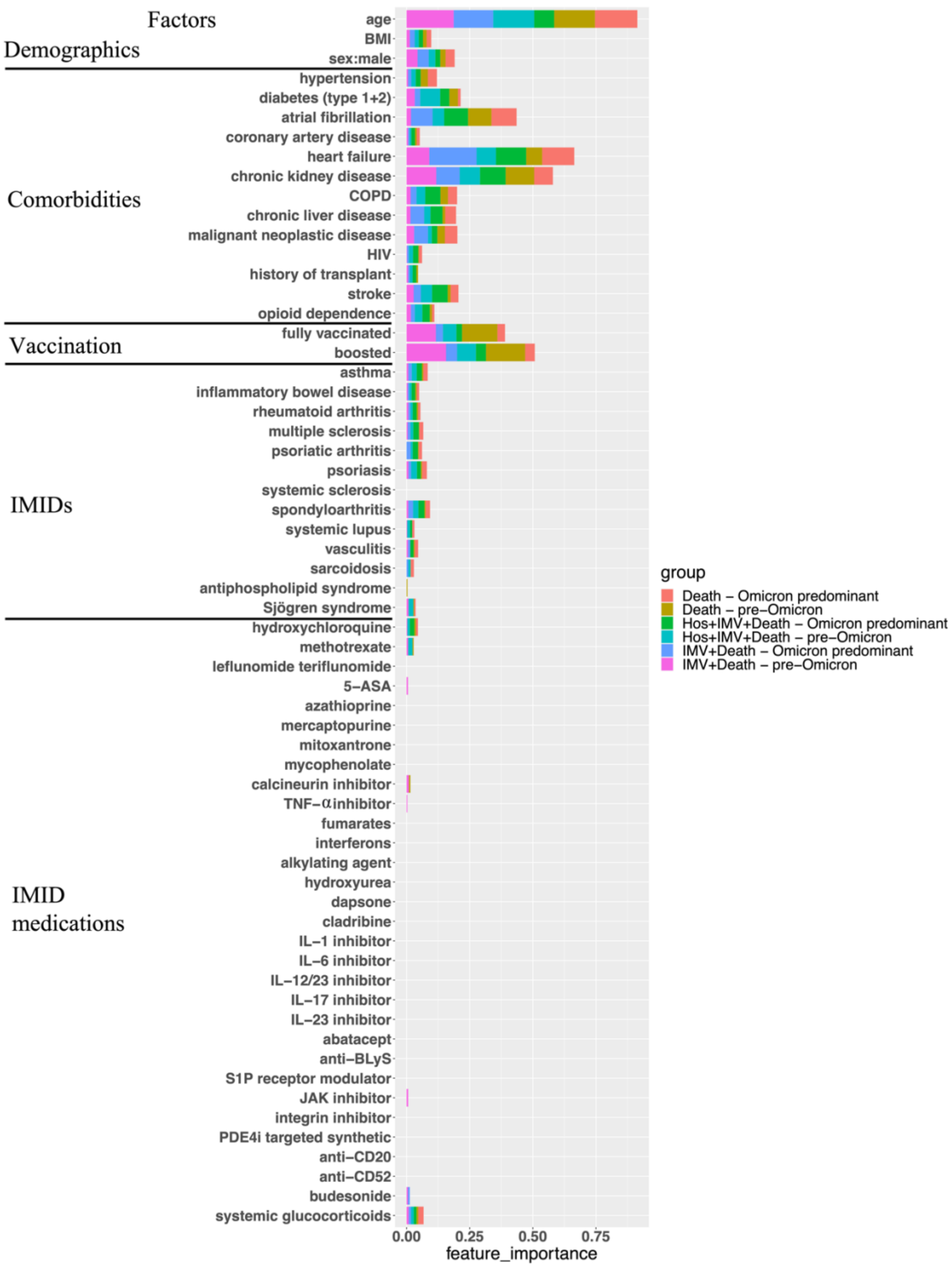

**Supplementary Figure S2. Feature importance for all extreme gradient boosting models**

Feature importance for all 63 factors for XGB models for hospitalisation and death across Time Interval 1 (01/03/2020 to 25/12/2021) and Time Interval 2 (26/12/2021 to 30/08/2022) using six different colours (coral, brown, green, cyan, blue, and purple). The feature importance of each feature is calculated using the number of times a feature is used to split the data across all trees.

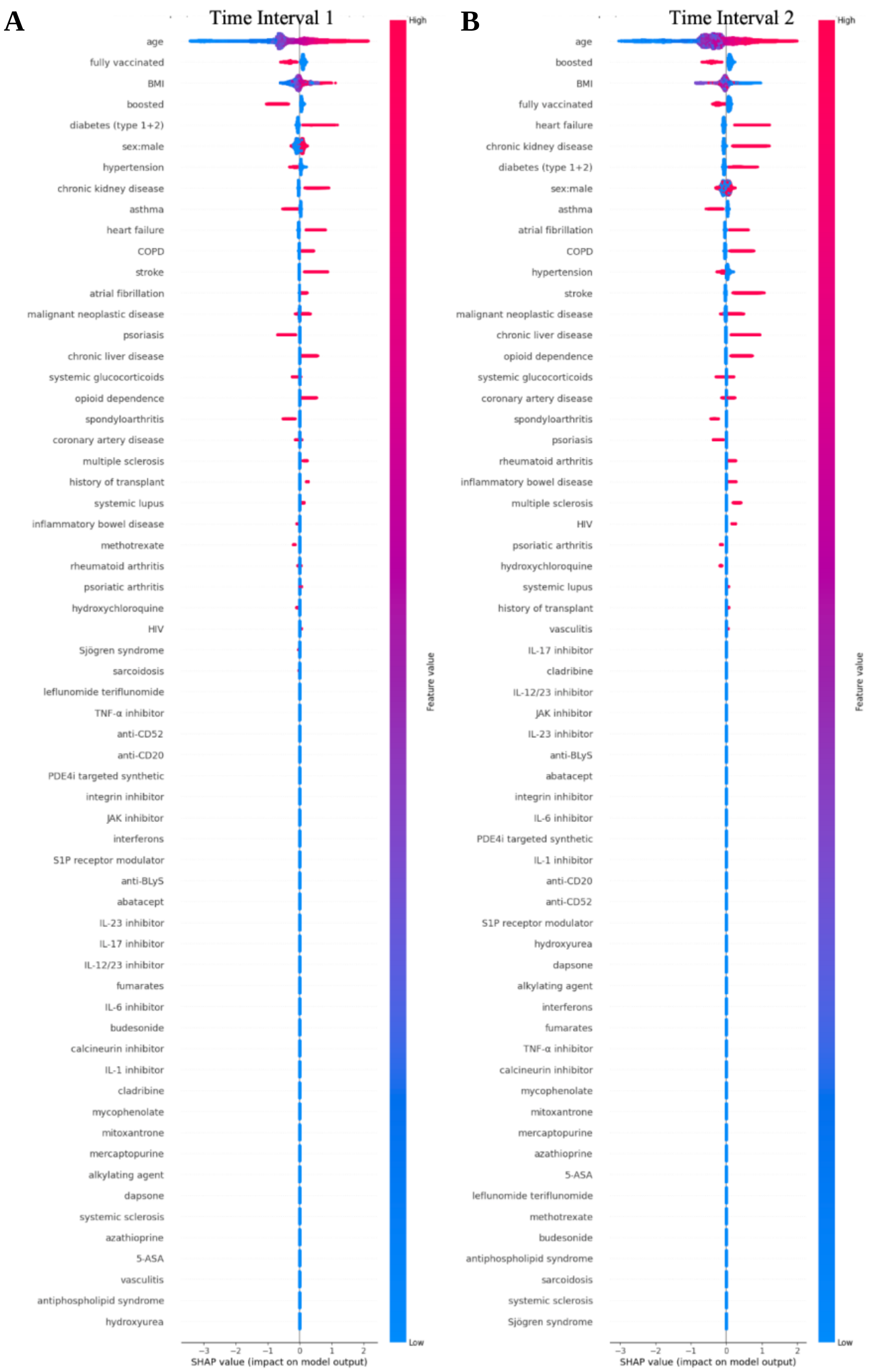

**Supplementary Figure S3. Feature importance for all features of all-age model of hospitalisation within the window of three days prior to 14 days of their first COVID-19 incidence, or MV or death within 30 days of their first COVID-19 incidence across Time Interval 1 (01/03/2020 to 25/12/2021) and Time Interval 2 (26/12/2021 to 30/08/2022).**

Extreme gradient boosting feature importance and influence of higher and lower values of the factors on the all-age group population outcome. The left side of each plot is associated with reduced risk and the right side of each plot is associated with increased risk. Red dots represent patients with higher values for a variable, and blue dots are patients with lower values. Nominal classes are binary [0, 1]. For sex, male is 1 (red).

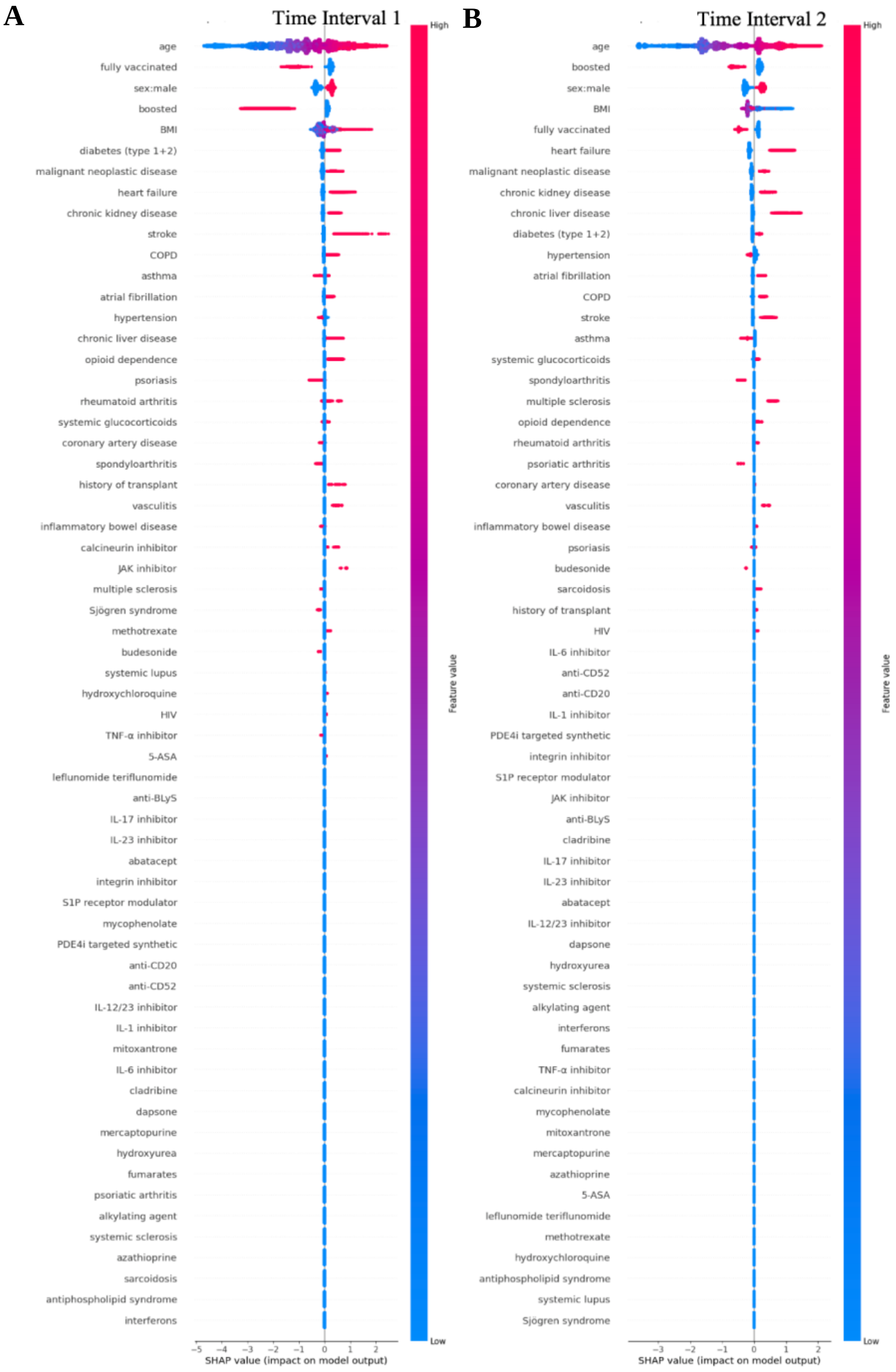

**Supplementary Figure S4. Feature importance for all features of all-age model of MV or death within 30 days of their first COVID-19 incidence across Time Interval 1 (01/03/2020 to 25/12/2021) and Time Interval 2 (26/12/2021 to 30/08/2022).**

Extreme gradient boosting feature importance and influence of higher and lower values of the factors on the all-age group population outcome. In both A and B plots, SHAP values < 0 are associated with reduced risk for mechanical ventilation or death, and SHAP values > 0 are associated with increased risk. Red dots represent patients with higher values for a variable, and blue dots are patients with lower values· Nominal classes are binary [0, 1]. For sex, male is 1 (red).

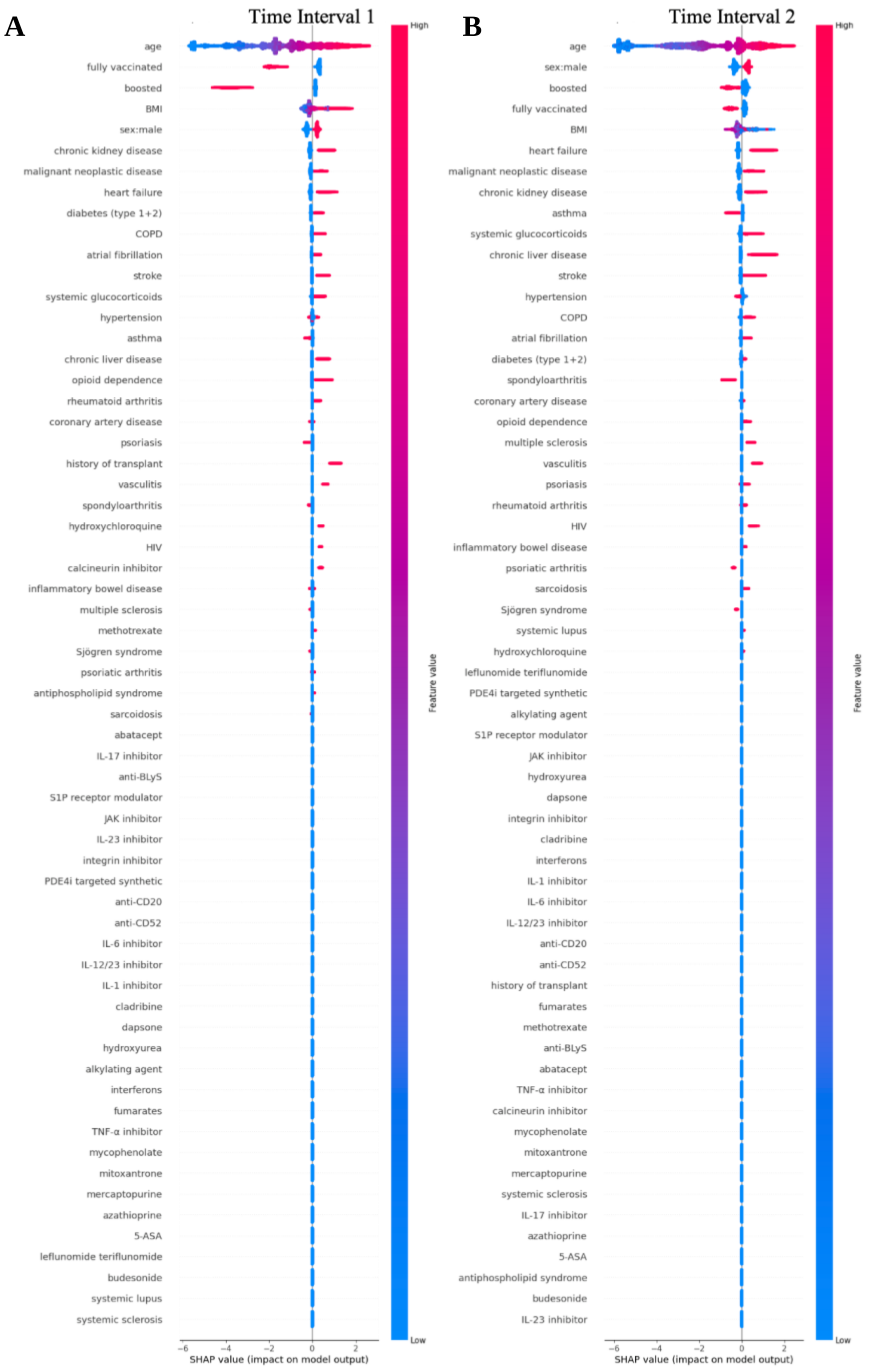

**Supplementary Figure S5. Feature importance for all features of all-age model of death within 30 days of their first COVID-19 incidence across Time Interval 1 (01/03/2020 to 25/12/2021) and Time Interval 2 (26/12/2021 to 30/08/2022).**

Extreme gradient boosting feature importance and influence of higher and lower values of the factors on the all-age group population outcome. In both A and B plots, SHAP values < 0 are associated with reduced risk for mechanical ventilation or death, and SHAP values > 0 are associated with increased risk. Red dots represent patients with higher values for a variable, and blue dots are patients with lower values. Nominal classes are binary [0, 1]. For sex, male is 1 (red).

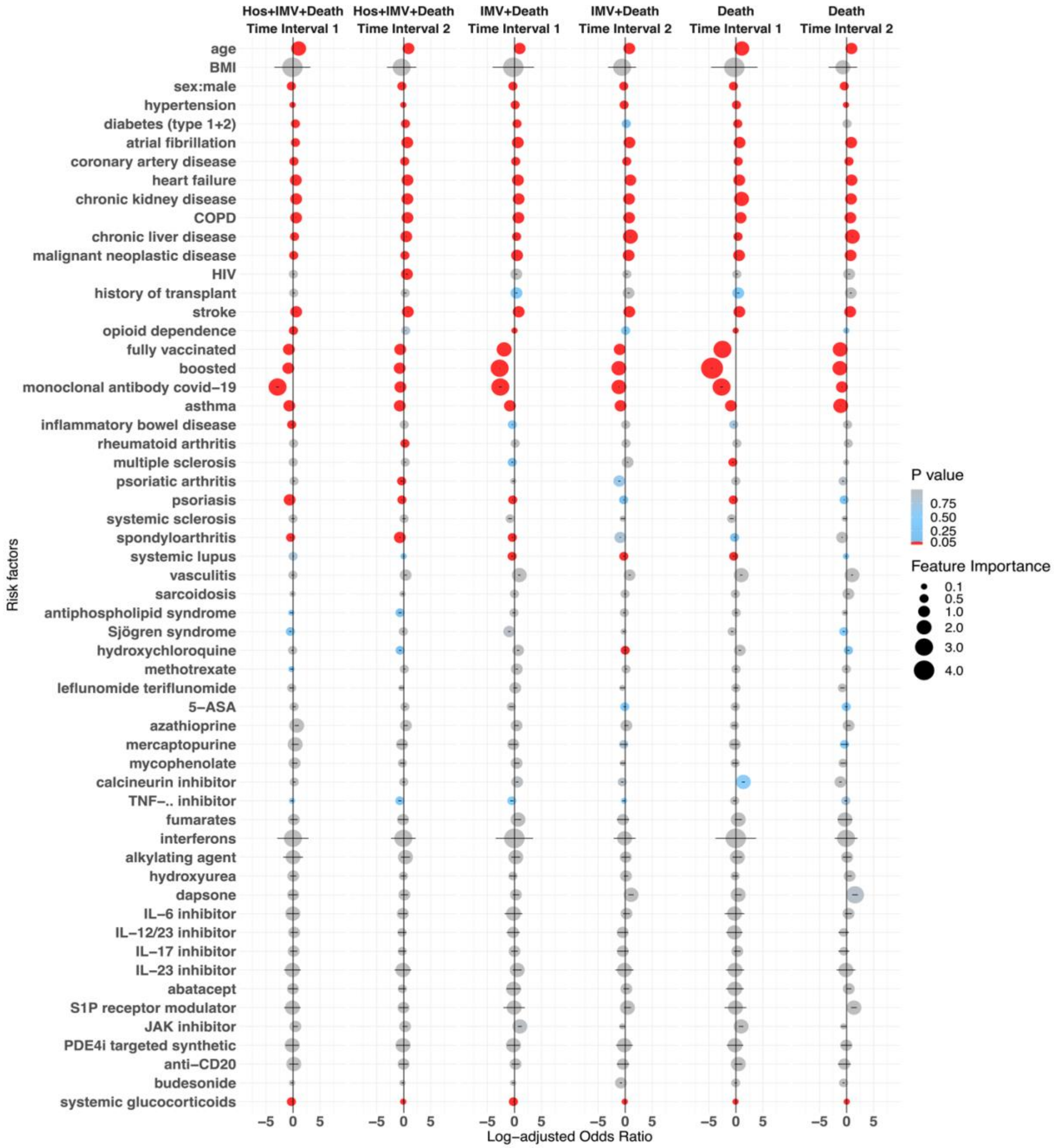

**Supplementary Figure S6. Log transformed adjusted odds ratio of logistic regression models, including anti-SARS-CoV-2 neutralizing monoclonal antibody factor.**

Selected factors for hospitalisation, MV or death, MV or death, and death across Time Interval 1 (01/03/2020 to 25/12/2021) and Time Interval 2 (26/12/2021 to 30/08/2022) in COVID-19 positive patients, using multivariable logistic regression (LR). The colour of dots represent P-value calculated using raw data. Position and size of dots represents log-adjusted odds ratio and feature importance from over-sampling. The error bar represents the 95% confidence interval. Factors with fewer than ten observations were excluded.

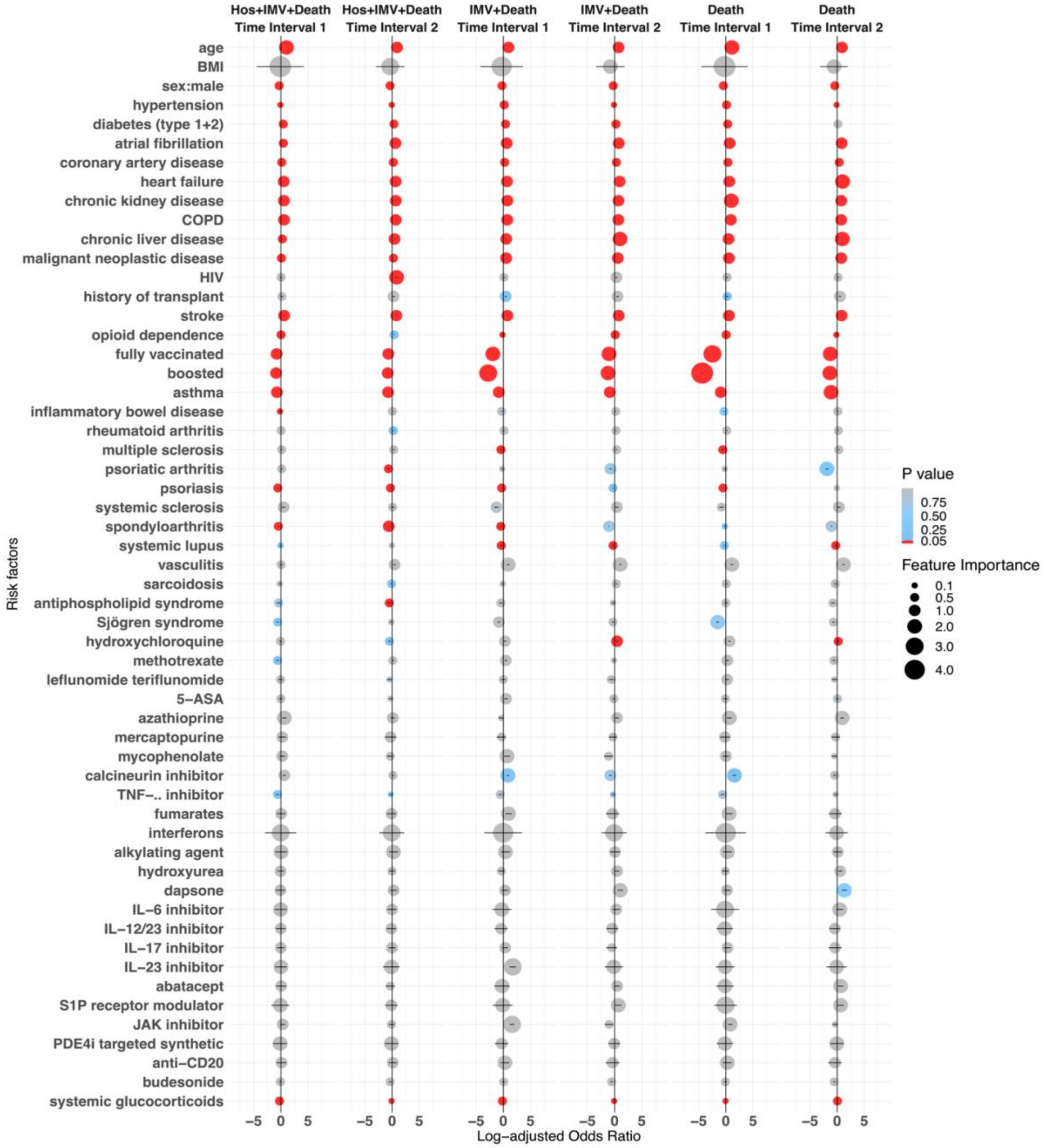

**Supplementary Figure S7. Log transformed adjusted odds ratio of logistic regression models, excluding patients treated with anti-SARS-CoV-2 neutralizing monoclonal antibodies.**

Selected factors for hospitalisation, MV or death, MV or death, and death across Time Interval 1 (01/03/2020 to 25/12/2021) and Time Interval 2 (26/12/2021 to 30/08/2022) in COVID-19 positive patients, using multivariable logistic regression (LR). The colour of dots represent P-value calculated using raw data. Position and size of dots represents log-adjusted odds ratio and feature importance from over-sampling. The error bar represents the 95% confidence interval. Factors with fewer than ten observations were excluded.

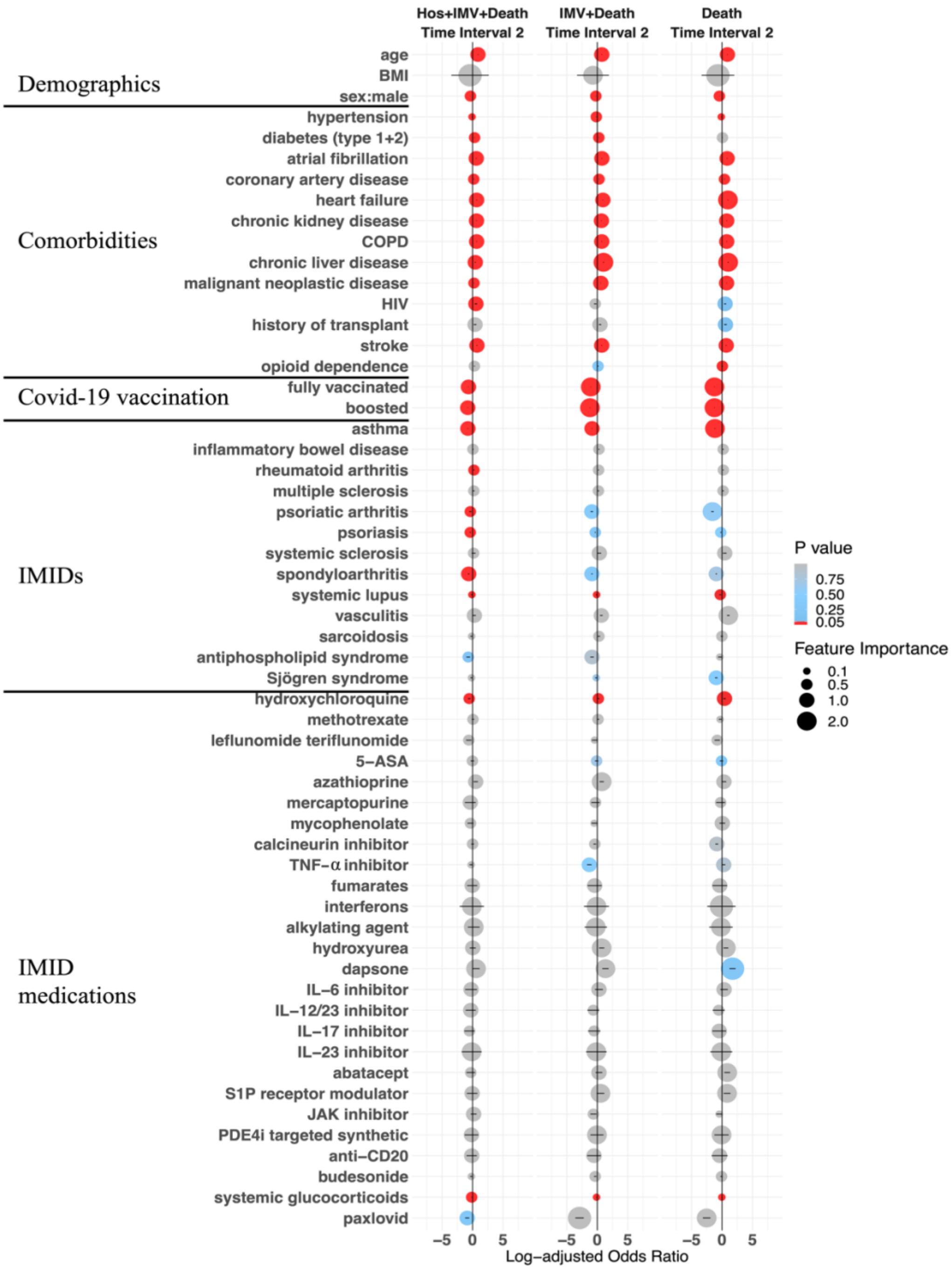

**Supplementary Figure S8. Log transformed adjusted odds ratio of logistic regression models, including the usage of paxlovid as an additional risk factor.**
Selected factors for hospitalisation, MV or death, MV or death, and death Time Interval 2 (26/12/2021 to 30/08/2022) in COVID-19 positive patients, using multivariable logistic regression (LR). The colour of dots represent P-value calculated using raw data. Position and size of dots represents log-adjusted odds ratio and feature importance from over-sampling. The error bar represents the 95% confidence interval. Factors with fewer than ten observations were excluded.

### Supplementary Tables

**Supplementary Table S1. XGB classification algorithm hyperparameters and hyperparameter range used in random-search tuning.**

Iteration number set to 1500.

| Name | Description | Range |
|---|---|---|
| learning_rate | boosting learning rate | (0·01, 0·05, 0·1, 0·2) |
| n_estimators | number of trees to fit | (5, 10, 15, …, 200) |
| max_depth | maximum tree depth | (2, 3, 5, …, 15) |
| min_child_weight | minimum sum of instance weight needed in a child | (0, 1, 2, ⋯, 50) |
| subsample | sub-sample ratio of the training instance | (0·4, 0·5, 0·6, …, 1·0) |
| colsample_bytree | sub-sample ratio of columns when constructing each tree | (0·4, 0·5, 0·6, …, 1·0) |
| reg_alpha | coefficient of L1 regularization for the node weights | (0, 1, 2, 3, 4, 5) |
| reg_lambda | coefficient of L2 regularization for the node weights | (1, 2, 3, 4, 5) |
| scale_pos_weight | control the balance of positive and negative weights, useful for unbalanced classes· | sum(negative instances) / sum(positive instances) |
| importance_type | define tree model importance type | (‘weight’) |
| random_state | random number seed | (42) |

**Supplementary Table S2. Results of the multivariable logistic regression (LR) model, Selected factors for hospitalisation, MV, or death at Time Interval 1 (01/03/2020 to 25/12/2021) in COVID-19 positive patients.**

| Feature | Adjusted odds ratio (AOR) | 95% confidence interval (low) | 95% confidence interval (high) | Log of AOR | Feature importance | P value | P value (FDR corrected) |
|---|---|---|---|---|---|---|---|
| age | 2.776 | 2.702 | 2.852 | 1.021 | 1.048 | < 0.0001 | < 0.0001 |
| BMI | 0.888 | 0.032 | 24.754 | -0.118 | 3.209 | 0.920 | 1.000 |
| sex: male | 0.735 | 0.723 | 0.746 | -0.308 | 0.293 | < 0.0001 | < 0.0001 |
| hypertension | 0.902 | 0.882 | 0.924 | -0.103 | 0.079 | < 0.0001 | < 0.0001 |
| diabetes (type 1+2) | 1.543 | 1.502 | 1.586 | 0.434 | 0.461 | < 0.0001 | < 0.0001 |
| atrial fibrillation | 1.553 | 1.492 | 1.616 | 0.440 | 0.480 | < 0.0001 | < 0.0001 |
| coronary artery disease | 1.178 | 1.133 | 1.224 | 0.164 | 0.202 | < 0.0001 | < 0.0001 |
| heart failure | 1.678 | 1.610 | 1.749 | 0.518 | 0.559 | < 0.0001 | < 0.0001 |
| chronic kidney disease | 1.781 | 1.718 | 1.846 | 0.577 | 0.613 | < 0.0001 | < 0.0001 |
| COPD | 1.779 | 1.704 | 1.857 | 0.576 | 0.619 | < 0.0001 | < 0.0001 |
| chronic liver disease | 1.337 | 1.250 | 1.432 | 0.291 | 0.359 | < 0.0001 | < 0.0001 |
| malignant neoplastic disease | 1.127 | 1.095 | 1.160 | 0.120 | 0.148 | < 0.0001 | < 0.0001 |
| HIV | 1.107 | 0.914 | 1.340 | 0.101 | 0.293 | 0.920 | 1.000 |
| history of transplant | 1.180 | 0.944 | 1.476 | 0.165 | 0.389 | 0.201 | 1.000 |
| stroke | 1.780 | 1.682 | 1.885 | 0.577 | 0.634 | < 0.0001 | < 0.0001 |
| opioid dependence | 1.079 | 1.007 | 1.155 | 0.076 | 0.144 | 0.001 | 0.013 |
| fully vaccinated | 0.459 | 0.450 | 0.468 | -0.779 | 0.760 | < 0.0001 | < 0.0001 |
| boosted | 0.420 | 0.407 | 0.433 | -0.869 | 0.838 | < 0.0001 | < 0.0001 |
| asthma | 0.493 | 0.479 | 0.509 | -0.707 | 0.676 | < 0.0001 | < 0.0001 |
| inflammatory bowel disease | 0.776 | 0.702 | 0.857 | -0.254 | 0.154 | 0.001 | 0.013 |

| | | | | | | | |
|---|---|---|---|---|---|---|---|
| rheumatoid arthritis | 1.098 | 1.016 | 1.186 | 0.094 | 0.171 | 0.499 | 1.000 |
| multiple sclerosis | 1.044 | 0.914 | 1.192 | 0.043 | 0.176 | 0.589 | 1.000 |
| psoriatic arthritis | 1.367 | 1.156 | 1.616 | 0.312 | 0.480 | 0.455 | 1.000 |
| psoriasis | 0.519 | 0.478 | 0.564 | -0.656 | 0.572 | < 0.0001 | < 0.0001 |
| systemic sclerosis | 0.994 | 0.729 | 1.355 | -0.006 | 0.304 | 0.826 | 1.000 |
| spondyloarthritis | 0.642 | 0.583 | 0.707 | -0.444 | 0.347 | < 0.0001 | < 0.0001 |
| systemic lupus | 0.990 | 0.867 | 1.132 | -0.010 | 0.124 | 0.042 | 0.452 |
| vasculitis | 0.942 | 0.733 | 1.210 | -0.060 | 0.191 | 0.215 | 1.000 |
| sarcoidosis | 0.875 | 0.719 | 1.064 | -0.134 | 0.062 | 0.297 | 1.000 |
| antiphospholipid syndrome | 0.705 | 0.528 | 0.942 | -0.349 | 0.060 | 0.038 | 0.427 |
| Sjögren’s syndrome | 0.520 | 0.425 | 0.635 | -0.655 | 0.454 | 0.007 | 0.086 |
| hydroxychloroquine | 0.869 | 0.704 | 1.071 | -0.141 | 0.069 | 0.162 | 1.000 |
| methotrexate | 0.677 | 0.543 | 0.845 | -0.389 | 0.168 | 0.009 | 0.106 |
| leflunomide teriflunomide | 0.771 | 0.503 | 1.181 | -0.260 | 0.166 | 0.736 | 1.000 |
| 5-ASA | 1.227 | 0.906 | 1.660 | 0.204 | 0.507 | 0.806 | 1.000 |
| azathioprine | 1.980 | 1.340 | 2.924 | 0.683 | 1.073 | 0.155 | 1.000 |
| mercaptopurine | 1.464 | 0.773 | 2.773 | 0.381 | 1.020 | 0.896 | 1.000 |
| mycophenolate | 1.372 | 0.818 | 2.300 | 0.316 | 0.833 | 0.923 | 1.000 |
| calcineurin inhibitor | 1.340 | 1.037 | 1.733 | 0.293 | 0.550 | 0.879 | 1.000 |
| TNF-α inhibitor | 0.818 | 0.618 | 1.082 | -0.201 | 0.079 | 0.048 | 0.496 |
| fumarates | 1.137 | 0.568 | 2.277 | 0.128 | 0.823 | 0.928 | 1.000 |
| interferons | 0.970 | 0.054 | 17.532 | -0.030 | 2.864 | 0.994 | 1.000 |
| alkylating agent | 1.001 | 0.160 | 6.253 | 0.001 | 1.833 | 0.982 | 1.000 |
| hydroxyurea | 1.037 | 0.584 | 1.839 | 0.036 | 0.609 | 0.785 | 1.000 |
| dapsone | 1.018 | 0.433 | 2.392 | 0.018 | 0.872 | 0.970 | 1.000 |
| IL-6 inhibitor | 0.956 | 0.300 | 3.047 | -0.045 | 1.114 | 0.947 | 1.000 |
| IL-12/23 inhibitor | 1.208 | 0.598 | 2.440 | 0.189 | 0.892 | 0.940 | 1.000 |
| IL-17 inhibitor | 1.099 | 0.567 | 2.128 | 0.094 | 0.755 | 0.989 | 1.000 |
| IL-23 inhibitor | 0.893 | 0.193 | 4.129 | -0.113 | 1.418 | 0.940 | 1.000 |
| abatacept | 0.978 | 0.399 | 2.399 | -0.022 | 0.875 | 0.965 | 1.000 |
| S1P receptor modulator | 0.889 | 0.194 | 4.067 | -0.118 | 1.403 | 0.896 | 1.000 |
| JAK inhibitor | 1.519 | 0.969 | 2.380 | 0.418 | 0.867 | 0.376 | 1.000 |
| PDE4i targeted synthetic | 0.857 | 0.244 | 3.007 | -0.154 | 1.101 | 0.889 | 1.000 |
| anti-CD20 | 1.144 | 0.481 | 2.718 | 0.134 | 1.000 | 0.873 | 1.000 |
| budesonide | 0.808 | 0.640 | 1.020 | -0.213 | 0.020 | 0.344 | 1.000 |
| systemic glucocorticoids | 0.714 | 0.686 | 0.744 | -0.337 | 0.296 | < 0.001 | < 0.001 |

**Supplementary Table S3. Results of the multivariable logistic regression (LR) model, Selected factors for hospitalisation, MV, or death at Time Interval 2 (26/12/2021 to 30/08/2022) in COVID-19 positive patients.**

| Feature | Adjusted odds ratio (AOR) | 95% confidence interval (low) | 95% confidence interval (high) | Log of AOR | Feature importance | P value | P value (FDR corrected) |
|---|---|---|---|---|---|---|---|
| age | 2.357 | 2.277 | 2.440 | 0.857 | 0.892 | < 0.0001 | < 0.0001 |
| BMI | 0.698 | 0.047 | 10.340 | -0.359 | 2.336 | 0.597 | 1.000 |
| sex: male | 0.700 | 0.687 | 0.713 | -0.357 | 0.338 | < 0.0001 | < 0.0001 |
| hypertension | 0.919 | 0.895 | 0.944 | -0.084 | 0.058 | < 0.0001 | < 0.0001 |
| diabetes (type 1+2) | 1.373 | 1.332 | 1.416 | 0.317 | 0.348 | < 0.0001 | < 0.0001 |

| | | | | | | | |
|---|---|---|---|---|---|---|---|
| atrial fibrillation | 1.863 | 1.791 | 1.939 | 0.622 | 0.662 | < 0.0001 | < 0.0001 |
| coronary artery disease | 1.189 | 1.143 | 1.237 | 0.173 | 0.213 | < 0.0001 | < 0.0001 |
| heart failure | 1.977 | 1.898 | 2.059 | 0.682 | 0.722 | < 0.0001 | < 0.0001 |
| chronic kidney disease | 1.935 | 1.866 | 2.006 | 0.660 | 0.696 | < 0.0001 | < 0.0001 |
| COPD | 1.976 | 1.895 | 2.061 | 0.681 | 0.723 | < 0.0001 | < 0.0001 |
| chronic liver disease | 1.581 | 1.477 | 1.694 | 0.458 | 0.527 | < 0.0001 | < 0.0001 |
| malignant neoplastic disease | 1.235 | 1.198 | 1.273 | 0.211 | 0.241 | < 0.0001 | < 0.0001 |
| HIV | 1.825 | 1.514 | 2.197 | 0.601 | 0.787 | < 0.0001 | < 0.0001 |
| history of transplant | 1.268 | 1.017 | 1.581 | 0.238 | 0.458 | 0.805 | 1.000 |
| stroke | 2.125 | 2.014 | 2.243 | 0.754 | 0.808 | < 0.0001 | < 0.0001 |
| opioid dependence | 1.433 | 1.338 | 1.536 | 0.360 | 0.429 | 0.064 | 0.636 |
| fully vaccinated | 0.516 | 0.504 | 0.528 | -0.662 | 0.639 | < 0.0001 | < 0.0001 |
| boosted | 0.467 | 0.454 | 0.479 | -0.762 | 0.736 | < 0.0001 | < 0.0001 |
| asthma | 0.476 | 0.460 | 0.492 | -0.743 | 0.710 | < 0.0001 | < 0.0001 |
| inflammatory bowel disease | 1.085 | 0.981 | 1.200 | 0.082 | 0.182 | 0.294 | 1.000 |
| rheumatoid arthritis | 1.241 | 1.143 | 1.347 | 0.216 | 0.298 | 0.006 | 0.067 |
| multiple sclerosis | 1.314 | 1.149 | 1.504 | 0.273 | 0.408 | 0.100 | 0.956 |
| psoriatic arthritis | 0.651 | 0.543 | 0.780 | -0.429 | 0.249 | 0.001 | 0.013 |
| psoriasis | 0.715 | 0.656 | 0.778 | -0.336 | 0.251 | < 0.0001 | < 0.0001 |
| systemic sclerosis | 1.064 | 0.776 | 1.459 | 0.062 | 0.378 | 0.681 | 1.000 |
| spondyloarthritis | 0.485 | 0.437 | 0.539 | -0.723 | 0.618 | < 0.0001 | < 0.0001 |
| systemic lupus | 0.944 | 0.826 | 1.079 | -0.057 | 0.076 | 0.005 | 0.061 |
| vasculitis | 1.391 | 1.107 | 1.745 | 0.330 | 0.557 | 0.473 | 1.000 |
| sarcoidosis | 0.769 | 0.622 | 0.950 | -0.263 | 0.051 | 0.124 | 1.000 |
| antiphospholipid syndrome | 0.523 | 0.389 | 0.703 | -0.648 | 0.352 | 0.007 | 0.075 |
| Sjögren’s syndrome | 0.916 | 0.760 | 1.104 | -0.088 | 0.099 | 0.537 | 1.000 |
| hydroxychloroquine | 0.515 | 0.423 | 0.628 | -0.663 | 0.465 | 0.006 | 0.067 |
| methotrexate | 1.087 | 0.888 | 1.330 | 0.083 | 0.285 | 0.845 | 1.000 |
| leflunomide teriflunomide | 0.673 | 0.446 | 1.017 | -0.396 | 0.017 | 0.397 | 1.000 |
| 5-ASA | 1.194 | 0.915 | 1.559 | 0.177 | 0.444 | 0.655 | 1.000 |
| azathioprine | 1.431 | 0.999 | 2.048 | 0.358 | 0.717 | 0.953 | 1.000 |
| mercaptopurine | 0.750 | 0.298 | 1.891 | -0.287 | 0.637 | 0.386 | 1.000 |
| mycophenolate | 0.757 | 0.465 | 1.234 | -0.278 | 0.210 | 0.490 | 1.000 |
| calcineurin inhibitor | 0.984 | 0.783 | 1.235 | -0.017 | 0.211 | 0.170 | 1.000 |
| TNF-α inhibitor | 0.497 | 0.369 | 0.670 | -0.699 | 0.401 | 0.025 | 0.258 |
| fumarates | 0.889 | 0.347 | 2.277 | -0.118 | 0.823 | 0.946 | 1.000 |
| interferons | 0.930 | 0.094 | 9.152 | -0.073 | 2.214 | 0.978 | 1.000 |
| alkylating agent | 1.269 | 0.540 | 2.986 | 0.239 | 1.094 | 0.672 | 1.000 |
| hydroxyurea | 0.948 | 0.564 | 1.594 | -0.054 | 0.466 | 0.915 | 1.000 |
| dapsone | 1.103 | 0.566 | 2.149 | 0.098 | 0.765 | 0.593 | 1.000 |
| IL-6 inhibitor | 0.850 | 0.361 | 2.004 | -0.163 | 0.695 | 0.857 | 1.000 |
| IL-12/23 inhibitor | 0.747 | 0.342 | 1.634 | -0.291 | 0.491 | 0.577 | 1.000 |
| IL-17 inhibitor | 0.775 | 0.387 | 1.550 | -0.256 | 0.438 | 0.693 | 1.000 |
| IL-23 inhibitor | 0.864 | 0.188 | 3.967 | -0.146 | 1.378 | 0.916 | 1.000 |
| abatacept | 0.786 | 0.400 | 1.545 | -0.241 | 0.435 | 0.780 | 1.000 |
| S1P receptor modulator | 0.951 | 0.397 | 2.280 | -0.050 | 0.824 | 0.983 | 1.000 |
| JAK inhibitor | 1.262 | 0.795 | 2.002 | 0.232 | 0.694 | 0.873 | 1.000 |

| | | | | | | | |
|---|---|---|---|---|---|---|---|
| PDE4i targeted synthetic | 0.875 | 0.250 | 3.071 | -0.133 | 1.122 | 0.935 | 1.000 |
| anti-CD20 | 0.970 | 0.405 | 2.323 | -0.030 | 0.843 | 0.988 | 1.000 |
| budesonide | 0.791 | 0.644 | 0.970 | -0.235 | 0.030 | 0.211 | 1.000 |
| systemic glucocorticoids | 0.9 | 0.866 | 0.934 | -0.106 | 0.068 | < 0.0001 | < 0.0001 |

**Supplementary Table S4. Results of the multivariable logistic regression (LR) model, Selected factors for MV or death at Time Interval 1 (01/03/2020 to 25/12/2021) in COVID-19 positive patients.**

| Feature | Adjusted odds ratio (AOR) | 95% confidence interval (low) | 95% confidence interval (high) | Log of AOR | Feature importance | P value | P value (FDR corrected) |
|---|---|---|---|---|---|---|---|
| age | 2.582 | 2.519 | 2.649 | 0.949 | 0.974 | < 0.0001 | < 0.0001 |
| BMI | 0.752 | 0.017 | 33.988 | -0.285 | 3.526 | 0.887 | 1.000 |
| sex: male | 0.750 | 0.738 | 0.762 | -0.288 | 0.272 | < 0.0001 | < 0.0001 |
| hypertension | 1.130 | 1.104 | 1.157 | 0.123 | 0.146 | < 0.0001 | < 0.0001 |
| diabetes (type 1+2) | 1.586 | 1.545 | 1.629 | 0.461 | 0.488 | < 0.0001 | < 0.0001 |
| atrial fibrillation | 1.859 | 1.793 | 1.927 | 0.620 | 0.656 | < 0.0001 | < 0.0001 |
| coronary artery disease | 1.247 | 1.203 | 1.293 | 0.221 | 0.257 | < 0.0001 | < 0.0001 |
| heart failure | 1.897 | 1.828 | 1.968 | 0.640 | 0.677 | < 0.0001 | < 0.0001 |
| chronic kidney disease | 2.139 | 2.069 | 2.210 | 0.760 | 0.793 | < 0.0001 | < 0.0001 |
| COPD | 2.043 | 1.964 | 2.125 | 0.715 | 0.754 | < 0.0001 | < 0.0001 |
| chronic liver disease | 1.492 | 1.398 | 1.590 | 0.400 | 0.464 | 0.002 | 0.026 |
| malignant neoplastic disease | 1.601 | 1.557 | 1.645 | 0.471 | 0.498 | < 0.0001 | < 0.0001 |
| HIV | 1.451 | 1.194 | 1.765 | 0.372 | 0.568 | 0.856 | 1.000 |
| history of transplant | 1.469 | 1.207 | 1.786 | 0.384 | 0.580 | 0.013 | 0.153 |
| stroke | 2.092 | 1.988 | 2.201 | 0.738 | 0.789 | < 0.0001 | < 0.0001 |
| opioid dependence | 1.012 | 0.946 | 1.082 | 0.012 | 0.079 | < 0.0001 | < 0.0001 |
| fully vaccinated | 0.145 | 0.142 | 0.149 | -1.929 | 1.906 | < 0.0001 | < 0.0001 |
| boosted | 0.064 | 0.061 | 0.067 | -2.752 | 2.704 | < 0.0001 | < 0.0001 |
| asthma | 0.428 | 0.414 | 0.442 | -0.849 | 0.817 | < 0.0001 | < 0.0001 |
| inflammatory bowel disease | 0.657 | 0.593 | 0.729 | -0.420 | 0.316 | 0.029 | 0.312 |
| rheumatoid arthritis | 1.122 | 1.038 | 1.213 | 0.115 | 0.193 | 0.969 | 1.000 |
| multiple sclerosis | 0.638 | 0.549 | 0.742 | -0.449 | 0.298 | 0.005 | 0.061 |
| psoriatic arthritis | 0.808 | 0.677 | 0.965 | -0.213 | 0.036 | 0.906 | 1.000 |
| psoriasis | 0.748 | 0.689 | 0.813 | -0.290 | 0.207 | < 0.0001 | < 0.0001 |
| systemic sclerosis | 0.373 | 0.259 | 0.538 | -0.986 | 0.620 | 0.100 | 0.890 |
| spondyloarthritis | 0.670 | 0.607 | 0.739 | -0.401 | 0.302 | 0.001 | 0.014 |
| systemic lupus | 0.633 | 0.550 | 0.728 | -0.458 | 0.317 | 0.001 | 0.014 |
| vasculitis | 2.654 | 2.113 | 3.333 | 0.976 | 1.204 | 0.233 | 1.000 |
| sarcoidosis | 1.005 | 0.829 | 1.219 | 0.005 | 0.198 | 0.313 | 1.000 |
| antiphospholipid syndrome | 0.891 | 0.677 | 1.172 | -0.115 | 0.159 | 0.944 | 1.000 |
| Sjögren’s syndrome | 0.344 | 0.277 | 0.429 | -1.066 | 0.847 | 0.070 | 0.670 |
| hydroxychloroquine | 2.132 | 1.719 | 2.643 | 0.757 | 0.972 | 0.383 | 1.000 |
| methotrexate | 1.530 | 1.236 | 1.893 | 0.425 | 0.638 | 0.293 | 1.000 |
| leflunomide teriflunomide | 1.200 | 0.834 | 1.726 | 0.182 | 0.546 | 0.181 | 1.000 |

| | | | | | | | |
|---|---|---|---|---|---|---|---|
| 5-ASA | 0.499 | 0.322 | 0.773 | -0.695 | 0.258 | 0.212 | 1.000 |
| azathioprine | 1.604 | 1.074 | 2.396 | 0.473 | 0.874 | 0.417 | 1.000 |
| mercaptopurine | 0.772 | 0.333 | 1.790 | -0.258 | 0.582 | 0.096 | 0.885 |
| mycophenolate | 1.603 | 0.944 | 2.721 | 0.472 | 1.001 | 0.967 | 1.000 |
| calcineurin inhibitor | 1.723 | 1.342 | 2.214 | 0.544 | 0.795 | 0.048 | 0.477 |
| TNF-α inhibitor | 0.543 | 0.399 | 0.739 | -0.610 | 0.302 | 0.023 | 0.258 |
| fumarates | 2.244 | 1.256 | 4.011 | 0.808 | 1.389 | 0.486 | 1.000 |
| interferons | 0.974 | 0.030 | 31.817 | -0.026 | 3.460 | 0.999 | 1.000 |
| alkylating agent | 1.342 | 0.589 | 3.059 | 0.294 | 1.118 | 0.825 | 1.000 |
| hydroxyurea | 0.699 | 0.365 | 1.338 | -0.358 | 0.291 | 0.741 | 1.000 |
| dapsone | 1.389 | 0.749 | 2.573 | 0.328 | 0.945 | 0.673 | 1.000 |
| IL-6 inhibitor | 0.797 | 0.149 | 4.255 | -0.227 | 1.448 | 0.619 | 1.000 |
| IL-12/23 inhibitor | 0.738 | 0.222 | 2.455 | -0.304 | 0.898 | 0.826 | 1.000 |
| IL-17 inhibitor | 0.988 | 0.532 | 1.833 | -0.012 | 0.606 | 0.796 | 1.000 |
| IL-23 inhibitor | 1.899 | 0.841 | 4.289 | 0.641 | 1.456 | 0.831 | 1.000 |
| abatacept | 0.807 | 0.215 | 3.031 | -0.215 | 1.109 | 0.981 | 1.000 |
| S1P receptor modulator | 0.928 | 0.124 | 6.931 | -0.075 | 1.936 | 0.985 | 1.000 |
| JAK inhibitor | 3.290 | 2.197 | 4.928 | 1.191 | 1.595 | 0.043 | 0.444 |
| PDE4i targeted synthetic | 0.783 | 0.232 | 2.646 | -0.244 | 0.973 | 0.867 | 1.000 |
| anti-CD20 | 1.281 | 0.616 | 2.664 | 0.247 | 0.980 | 0.845 | 1.000 |
| budesonide | 0.756 | 0.608 | 0.940 | -0.279 | 0.062 | 0.659 | 1.000 |
| systemic glucocorticoids | 0.818 | 0.786 | 0.852 | -0.200 | 0.160 | < 0.0001 | < 0.0001 |

**Supplementary Table S5. Results of the multivariable logistic regression (LR) model, Selected factors for MV or death at Time Interval 2 (26/12/2021 to 30/08/2022) in COVID-19 positive patients.**

| Feature | Adjusted odds ratio (AOR) | 95% confidence interval (low) | 95% confidence interval (high) | Log of AOR | Feature importance | P value | P value (FDR corrected) |
|---|---|---|---|---|---|---|---|
| age | 2.078 | 2.012 | 2.145 | 0.731 | 0.763 | < 0.0001 | < 0.0001 |
| BMI | 0.461 | 0.033 | 6.475 | -0.774 | 1.868 | 0.392 | 1.000 |
| sex: male | 0.767 | 0.754 | 0.781 | -0.265 | 0.247 | < 0.0001 | < 0.0001 |
| hypertension | 0.819 | 0.799 | 0.840 | -0.200 | 0.174 | < 0.0001 | < 0.0001 |
| diabetes (type 1+2) | 1.238 | 1.202 | 1.274 | 0.213 | 0.242 | 0.003 | 0.043 |
| atrial fibrillation | 2.151 | 2.079 | 2.226 | 0.766 | 0.800 | < 0.0001 | < 0.0001 |
| coronary artery disease | 1.330 | 1.284 | 1.379 | 0.285 | 0.321 | < 0.0001 | < 0.0001 |
| heart failure | 2.563 | 2.474 | 2.654 | 0.941 | 0.976 | < 0.0001 | < 0.0001 |
| chronic kidney disease | 2.026 | 1.962 | 2.092 | 0.706 | 0.738 | < 0.0001 | < 0.0001 |
| COPD | 2.040 | 1.964 | 2.117 | 0.713 | 0.750 | < 0.0001 | < 0.0001 |
| chronic liver disease | 2.562 | 2.413 | 2.721 | 0.941 | 1.001 | < 0.0001 | < 0.0001 |
| malignant neoplastic disease | 1.856 | 1.806 | 1.908 | 0.619 | 0.646 | < 0.0001 | < 0.0001 |
| HIV | 1.272 | 1.069 | 1.513 | 0.241 | 0.414 | 0.271 | 1.000 |
| history of transplant | 1.821 | 1.495 | 2.217 | 0.599 | 0.796 | 0.806 | 1.000 |
| stroke | 2.166 | 2.067 | 2.270 | 0.773 | 0.820 | < 0.0001 | < 0.0001 |
| opioid dependence | 1.057 | 0.991 | 1.126 | 0.055 | 0.119 | 0.057 | 0.640 |
| fully vaccinated | 0.364 | 0.356 | 0.373 | -1.010 | 0.986 | < 0.0001 | < 0.0001 |
| boosted | 0.306 | 0.298 | 0.314 | -1.185 | 1.159 | < 0.0001 | < 0.0001 |

| | | | | | | | |
|---|---|---|---|---|---|---|---|
| asthma | 0.424 | 0.410 | 0.439 | -0.857 | 0.823 | < 0.0001 | < 0.0001 |
| inflammatory bowel disease | 1.064 | 0.968 | 1.170 | 0.062 | 0.157 | 0.481 | 1.000 |
| rheumatoid arthritis | 1.156 | 1.068 | 1.251 | 0.145 | 0.224 | 0.594 | 1.000 |
| multiple sclerosis | 1.548 | 1.370 | 1.751 | 0.437 | 0.560 | 0.397 | 1.000 |
| psoriatic arthritis | 0.390 | 0.319 | 0.477 | -0.941 | 0.740 | 0.062 | 0.667 |
| psoriasis | 0.802 | 0.738 | 0.871 | -0.221 | 0.138 | 0.003 | 0.043 |
| systemic sclerosis | 0.581 | 0.405 | 0.834 | -0.543 | 0.182 | 0.428 | 1.000 |
| spondyloarthritis | 0.392 | 0.353 | 0.435 | -0.936 | 0.832 | 0.047 | 0.552 |
| systemic lupus | 0.763 | 0.668 | 0.871 | -0.271 | 0.138 | 0.001 | 0.016 |
| vasculitis | 1.989 | 1.649 | 2.399 | 0.688 | 0.875 | 0.749 | 1.000 |
| sarcoidosis | 0.969 | 0.796 | 1.181 | -0.031 | 0.166 | 0.924 | 1.000 |
| antiphospholipid syndrome | 0.901 | 0.697 | 1.165 | -0.104 | 0.153 | 0.217 | 1.000 |
| Sjögren's syndrome | 0.807 | 0.665 | 0.981 | -0.214 | 0.019 | 0.089 | 0.851 |
| hydroxychloroquine | 1.036 | 0.870 | 1.232 | 0.035 | 0.209 | 0.005 | 0.068 |
| methotrexate | 1.039 | 0.859 | 1.256 | 0.038 | 0.228 | 0.335 | 1.000 |
| leflunomide teriflunomide | 0.512 | 0.332 | 0.791 | -0.670 | 0.235 | 0.412 | 1.000 |
| 5-ASA | 0.982 | 0.753 | 1.281 | -0.018 | 0.248 | 0.025 | 0.323 |
| azathioprine | 1.323 | 0.918 | 1.908 | 0.280 | 0.646 | 0.468 | 1.000 |
| mercaptopurine | 0.713 | 0.354 | 1.438 | -0.338 | 0.363 | 0.079 | 0.785 |
| mycophenolate | 0.574 | 0.354 | 0.931 | -0.555 | 0.071 | 0.815 | 1.000 |
| calcineurin inhibitor | 0.562 | 0.452 | 0.699 | -0.576 | 0.358 | 0.078 | 0.785 |
| TNF- α inhibitor | 0.805 | 0.603 | 1.075 | -0.217 | 0.072 | 0.038 | 0.467 |
| fumarates | 0.587 | 0.190 | 1.820 | -0.532 | 0.599 | 0.724 | 1.000 |
| interferons | 0.856 | 0.113 | 6.488 | -0.155 | 1.870 | 0.997 | 1.000 |
| alkylating agent | 1.131 | 0.502 | 2.550 | 0.123 | 0.936 | 0.732 | 1.000 |
| hydroxyurea | 1.240 | 0.823 | 1.868 | 0.215 | 0.625 | 0.630 | 1.000 |
| dapsone | 3.975 | 2.340 | 6.753 | 1.380 | 1.910 | 0.223 | 1.000 |
| IL-6 inhibitor | 1.407 | 0.754 | 2.627 | 0.342 | 0.966 | 0.705 | 1.000 |
| IL-12/23 inhibitor | 0.527 | 0.194 | 1.429 | -0.641 | 0.357 | 0.668 | 1.000 |
| IL-17 inhibitor | 0.566 | 0.200 | 1.602 | -0.570 | 0.471 | 0.695 | 1.000 |
| IL-23 inhibitor | 0.843 | 0.159 | 4.473 | -0.171 | 1.498 | 0.981 | 1.000 |
| abatacept | 1.371 | 0.805 | 2.335 | 0.316 | 0.848 | 0.778 | 1.000 |
| S1P receptor modulator | 1.735 | 0.908 | 3.317 | 0.551 | 1.199 | 0.694 | 1.000 |
| JAK inhibitor | 0.533 | 0.323 | 0.877 | -0.630 | 0.131 | 0.862 | 1.000 |
| PDE4i targeted synthetic | 0.809 | 0.182 | 3.593 | -0.211 | 1.279 | 0.984 | 1.000 |
| anti-CD20 | 0.568 | 0.185 | 1.737 | -0.566 | 0.552 | 0.915 | 1.000 |
| budesonide | 0.483 | 0.389 | 0.599 | -0.728 | 0.512 | 0.162 | 1.000 |
| systemic glucocorticoids | 0.921 | 0.889 | 0.954 | -0.083 | 0.047 | < 0.001 | < 0.001 |

**Supplementary Table S6. Results of the multivariable logistic regression (LR) model, Selected factors for death at Time Interval 1 (01/03/2020 to 25/12/2021) in COVID-19 positive patients.**

| Feature | Adjusted odds ratio (AOR) | 95% confidence interval (low) | 95% confidence interval (high) | Log of AOR | Feature importance | P value | P value (FDR corrected) |
|---|---|---|---|---|---|---|---|
| age | 2.951 | 2.878 | 3.025 | 1.082 | 1.107 | < 0.0001 | < 0.0001 |
| BMI | 0.786 | 0.011 | 56.374 | -0.240 | 4.032 | 0.919 | 1.000 |

| | | | | | | | |
|---|---|---|---|---|---|---|---|
| sex: male | 0.642 | 0.631 | 0.653 | -0.443 | 0.426 | < 0.0001 | < 0.0001 |
| hypertension | 1.115 | 1.089 | 1.142 | 0.109 | 0.133 | < 0.0001 | < 0.0001 |
| diabetes (type 1+2) | 1.398 | 1.359 | 1.438 | 0.335 | 0.363 | < 0.0001 | < 0.0001 |
| atrial fibrillation | 1.996 | 1.923 | 2.073 | 0.691 | 0.729 | < 0.0001 | < 0.0001 |
| coronary artery disease | 1.527 | 1.471 | 1.584 | 0.423 | 0.460 | < 0.0001 | < 0.0001 |
| heart failure | 1.865 | 1.793 | 1.939 | 0.623 | 0.662 | < 0.0001 | < 0.0001 |
| chronic kidney disease | 2.825 | 2.729 | 2.924 | 1.039 | 1.073 | < 0.0001 | < 0.0001 |
| COPD | 2.378 | 2.282 | 2.479 | 0.866 | 0.908 | < 0.0001 | < 0.0001 |
| chronic liver disease | 1.449 | 1.354 | 1.551 | 0.371 | 0.439 | 0.002 | 0.027 |
| malignant neoplastic disease | 1.783 | 1.732 | 1.837 | 0.579 | 0.608 | < 0.0001 | < 0.0001 |
| HIV | 1.221 | 0.997 | 1.493 | 0.199 | 0.401 | 0.605 | 1.000 |
| history of transplant | 1.654 | 1.326 | 2.063 | 0.503 | 0.724 | 0.017 | 0.200 |
| stroke | 1.892 | 1.793 | 1.996 | 0.638 | 0.691 | < 0.0001 | < 0.0001 |
| opioid dependence | 0.968 | 0.902 | 1.038 | -0.033 | 0.037 | < 0.0001 | < 0.0001 |
| fully vaccinated | 0.082 | 0.080 | 0.085 | -2.498 | 2.470 | < 0.0001 | < 0.0001 |
| boosted | 0.012 | 0.011 | 0.013 | -4.415 | 4.329 | < 0.0001 | < 0.0001 |
| asthma | 0.395 | 0.383 | 0.409 | -0.928 | 0.895 | < 0.0001 | < 0.0001 |
| inflammatory bowel disease | 0.684 | 0.614 | 0.763 | -0.379 | 0.271 | 0.077 | 0.795 |
| rheumatoid arthritis | 1.191 | 1.097 | 1.292 | 0.175 | 0.256 | 0.345 | 1.000 |
| multiple sclerosis | 0.549 | 0.465 | 0.648 | -0.600 | 0.434 | < 0.0001 | < 0.0001 |
| psoriatic arthritis | 0.976 | 0.807 | 1.181 | -0.024 | 0.166 | 0.542 | 1.000 |
| psoriasis | 0.584 | 0.534 | 0.640 | -0.537 | 0.446 | < 0.0001 | < 0.0001 |
| systemic sclerosis | 0.508 | 0.349 | 0.740 | -0.677 | 0.301 | 0.322 | 1.000 |
| spondyloarthritis | 0.778 | 0.703 | 0.861 | -0.251 | 0.150 | 0.012 | 0.148 |
| systemic lupus | 0.693 | 0.602 | 0.799 | -0.367 | 0.225 | 0.005 | 0.065 |
| vasculitis | 2.753 | 2.186 | 3.469 | 1.013 | 1.244 | 0.640 | 1.000 |
| sarcoidosis | 0.904 | 0.739 | 1.107 | -0.101 | 0.102 | 0.873 | 1.000 |
| antiphospholipid syndrome | 1.078 | 0.815 | 1.426 | 0.075 | 0.355 | 0.748 | 1.000 |
| Sjögren’s syndrome | 0.492 | 0.393 | 0.615 | -0.710 | 0.486 | 0.346 | 1.000 |
| hydroxychloroquine | 2.142 | 1.721 | 2.667 | 0.762 | 0.981 | 0.664 | 1.000 |
| methotrexate | 1.028 | 0.796 | 1.326 | 0.027 | 0.282 | 0.641 | 1.000 |
| leflunomide teriflunomide | 1.003 | 0.654 | 1.537 | 0.003 | 0.430 | 0.373 | 1.000 |
| 5-ASA | 0.910 | 0.631 | 1.314 | -0.094 | 0.273 | 0.538 | 1.000 |
| azathioprine | 0.790 | 0.493 | 1.267 | -0.235 | 0.237 | 0.896 | 1.000 |
| mercaptopurine | 0.851 | 0.383 | 1.895 | -0.161 | 0.639 | 0.199 | 1.000 |
| mycophenolate | 0.948 | 0.524 | 1.716 | -0.053 | 0.540 | 0.793 | 1.000 |
| calcineurin inhibitor | 4.043 | 3.043 | 5.371 | 1.397 | 1.681 | 0.020 | 0.225 |
| TNF- α inhibitor | 0.829 | 0.600 | 1.145 | -0.188 | 0.135 | 0.076 | 0.795 |
| fumarates | 1.423 | 0.733 | 2.765 | 0.353 | 1.017 | 0.847 | 1.000 |
| interferons | 0.980 | 0.023 | 42.692 | -0.020 | 3.754 | 1.000 | 1.000 |
| alkylating agent | 1.253 | 0.524 | 2.998 | 0.226 | 1.098 | 0.826 | 1.000 |
| hydroxyurea | 0.893 | 0.513 | 1.553 | -0.114 | 0.440 | 0.345 | 1.000 |
| dapsone | 1.416 | 0.771 | 2.601 | 0.348 | 0.956 | 0.443 | 1.000 |
| IL-6 inhibitor | 0.815 | 0.133 | 5.013 | -0.205 | 1.612 | 0.622 | 1.000 |
| IL-12/23 inhibitor | 0.798 | 0.182 | 3.501 | -0.226 | 1.253 | 0.900 | 1.000 |
| IL-17 inhibitor | 1.208 | 0.646 | 2.259 | 0.189 | 0.815 | 0.899 | 1.000 |
| IL-23 inhibitor | 0.880 | 0.160 | 4.840 | -0.128 | 1.577 | 0.984 | 1.000 |
| abatacept | 0.854 | 0.168 | 4.349 | -0.157 | 1.470 | 0.989 | 1.000 |

| | | | | | | | |
|---|---|---|---|---|---|---|---|
| S1P receptor modulator | 0.931 | 0.122 | 7.092 | -0.072 | 1.959 | 0.973 | 1.000 |
| JAK inhibitor | 2.283 | 1.501 | 3.473 | 0.826 | 1.245 | 0.109 | 1.000 |
| PDE4i targeted synthetic | 0.874 | 0.187 | 4.092 | -0.134 | 1.409 | 0.968 | 1.000 |
| anti-CD20 | 1.444 | 0.723 | 2.881 | 0.367 | 1.058 | 0.837 | 1.000 |
| budesonide | 0.948 | 0.751 | 1.197 | -0.054 | 0.180 | 0.314 | 1.000 |
| systemic glucocorticoids | 0.934 | 0.897 | 0.974 | -0.068 | 0.026 | < 0.001 | < 0.001 |

**Supplementary Table S7. Results of the multivariable logistic regression (LR) model, Selected factors for death at Time Interval 2 (26/12/2021 to 30/08/2022) in COVID-19 positive patients.**

| Feature | Adjusted odds ratio (AOR) | 95% confidence interval (low) | 95% confidence interval (high) | Log of AOR | Feature importance | P value | P value (FDR corrected) |
|---|---|---|---|---|---|---|---|
| age | 2.474 | 2.396 | 2.555 | 0.906 | 0.938 | < 0.0001 | < 0.0001 |
| BMI | 0.563 | 0.042 | 7.614 | -0.575 | 2.030 | 0.384 | 1.000 |
| sex: male | 0.646 | 0.634 | 0.658 | -0.437 | 0.418 | < 0.0001 | < 0.0001 |
| hypertension | 0.883 | 0.861 | 0.906 | -0.125 | 0.099 | < 0.0001 | < 0.0001 |
| diabetes (type 1+2) | 1.085 | 1.053 | 1.117 | 0.081 | 0.111 | 0.244 | 1.000 |
| atrial fibrillation | 2.251 | 2.175 | 2.330 | 0.812 | 0.846 | < 0.0001 | < 0.0001 |
| coronary artery disease | 1.537 | 1.484 | 1.594 | 0.430 | 0.466 | < 0.0001 | < 0.0001 |
| heart failure | 2.479 | 2.392 | 2.568 | 0.908 | 0.943 | < 0.0001 | < 0.0001 |
| chronic kidney disease | 2.223 | 2.151 | 2.296 | 0.799 | 0.831 | < 0.0001 | < 0.0001 |
| COPD | 2.009 | 1.935 | 2.088 | 0.698 | 0.736 | < 0.0001 | < 0.0001 |
| chronic liver disease | 2.796 | 2.635 | 2.968 | 1.028 | 1.088 | < 0.0001 | < 0.0001 |
| malignant neoplastic disease | 2.084 | 2.026 | 2.143 | 0.734 | 0.762 | < 0.0001 | < 0.0001 |
| HIV | 1.634 | 1.381 | 1.933 | 0.491 | 0.659 | 0.130 | 1.000 |
| history of transplant | 2.031 | 1.674 | 2.465 | 0.709 | 0.902 | 0.337 | 1.000 |
| stroke | 1.870 | 1.784 | 1.960 | 0.626 | 0.673 | < 0.0001 | < 0.0001 |
| opioid dependence | 0.946 | 0.883 | 1.012 | -0.056 | 0.012 | 0.001 | 0.016 |
| fully vaccinated | 0.301 | 0.294 | 0.308 | -1.201 | 1.177 | < 0.0001 | < 0.0001 |
| boosted | 0.286 | 0.278 | 0.293 | -1.253 | 1.227 | < 0.0001 | < 0.0001 |
| asthma | 0.331 | 0.319 | 0.343 | -1.106 | 1.070 | < 0.0001 | < 0.0001 |
| inflammatory bowel disease | 1.078 | 0.983 | 1.183 | 0.076 | 0.168 | 0.905 | 1.000 |
| rheumatoid arthritis | 1.316 | 1.218 | 1.422 | 0.275 | 0.352 | 0.273 | 1.000 |
| multiple sclerosis | 0.923 | 0.805 | 1.060 | -0.080 | 0.058 | 0.526 | 1.000 |
| psoriatic arthritis | 0.519 | 0.432 | 0.623 | -0.657 | 0.474 | 0.084 | 0.803 |
| psoriasis | 0.647 | 0.593 | 0.705 | -0.436 | 0.350 | 0.003 | 0.046 |
| systemic sclerosis | 0.746 | 0.535 | 1.040 | -0.293 | 0.039 | 0.381 | 1.000 |
| spondyloarthritis | 0.446 | 0.401 | 0.496 | -0.807 | 0.701 | 0.054 | 0.581 |
| systemic lupus | 0.872 | 0.766 | 0.993 | -0.137 | 0.007 | 0.011 | 0.150 |
| vasculitis | 2.640 | 2.181 | 3.193 | 0.971 | 1.161 | 0.927 | 1.000 |
| sarcoidosis | 1.395 | 1.155 | 1.685 | 0.333 | 0.522 | 0.440 | 1.000 |
| antiphospholipid syndrome | 0.746 | 0.578 | 0.964 | -0.293 | 0.037 | 0.096 | 0.885 |
| Sjögren’s syndrome | 0.607 | 0.500 | 0.738 | -0.499 | 0.304 | 0.022 | 0.271 |
| hydroxychloroquine | 1.401 | 1.183 | 1.659 | 0.337 | 0.506 | 0.004 | 0.057 |
| methotrexate | 0.911 | 0.750 | 1.107 | -0.093 | 0.102 | 0.200 | 1.000 |

| | | | | | | | |
|---|---|---|---|---|---|---|---|
| leflunomide teriflunomide | 0.502 | 0.332 | 0.758 | -0.690 | 0.277 | 0.472 | 1.000 |
| 5-ASA | 0.992 | 0.760 | 1.296 | -0.008 | 0.259 | 0.013 | 0.168 |
| azathioprine | 1.417 | 0.998 | 2.012 | 0.348 | 0.699 | 0.384 | 1.000 |
| mercaptopurine | 0.732 | 0.352 | 1.527 | -0.311 | 0.423 | 0.039 | 0.458 |
| mycophenolate | 0.553 | 0.335 | 0.912 | -0.593 | 0.092 | 0.805 | 1.000 |
| calcineurin inhibitor | 0.351 | 0.279 | 0.441 | -1.047 | 0.819 | 0.075 | 0.745 |
| TNF- α inhibitor | 0.874 | 0.664 | 1.151 | -0.134 | 0.141 | 0.049 | 0.550 |
| fumarates | 0.780 | 0.207 | 2.942 | -0.249 | 1.079 | 0.802 | 1.000 |
| interferons | 0.923 | 0.113 | 7.561 | -0.081 | 2.023 | 0.999 | 1.000 |
| alkylating agent | 1.080 | 0.502 | 2.326 | 0.077 | 0.844 | 0.654 | 1.000 |
| hydroxyurea | 1.655 | 1.124 | 2.438 | 0.504 | 0.891 | 0.495 | 1.000 |
| dapsone | 3.786 | 2.333 | 6.147 | 1.331 | 1.816 | 0.072 | 0.744 |
| IL-6 inhibitor | 1.359 | 0.717 | 2.573 | 0.307 | 0.945 | 0.617 | 1.000 |
| IL-12/23 inhibitor | 0.627 | 0.241 | 1.631 | -0.467 | 0.489 | 0.640 | 1.000 |
| IL-17 inhibitor | 0.650 | 0.243 | 1.740 | -0.431 | 0.554 | 0.730 | 1.000 |
| IL-23 inhibitor | 0.901 | 0.160 | 5.068 | -0.104 | 1.623 | 0.987 | 1.000 |
| abatacept | 1.460 | 0.901 | 2.363 | 0.378 | 0.860 | 0.708 | 1.000 |
| S1P receptor modulator | 3.246 | 1.944 | 5.419 | 1.178 | 1.690 | 0.587 | 1.000 |
| JAK inhibitor | 0.629 | 0.381 | 1.040 | -0.464 | 0.039 | 0.704 | 1.000 |
| PDE4i targeted synthetic | 0.939 | 0.345 | 2.560 | -0.063 | 0.940 | 0.774 | 1.000 |
| anti-CD20 | 0.699 | 0.218 | 2.246 | -0.358 | 0.809 | 0.947 | 1.000 |
| budesonide | 0.593 | 0.485 | 0.725 | -0.522 | 0.321 | 0.257 | 1.000 |
| systemic glucocorticoids | 1.020 | 0.984 | 1.058 | 0.020 | 0.056 | < 0.0001 | < 0.0001 |

**Supplementary Table S8. SNOMED definitions of IMIDs in this paper.**

The SNOMED Concept IDs used to define certain types of IMIDs in the research paper.

| IMID type | SNOMED Concept ID |
|---|---|
| asthma | 266361008, 426656000, 405944004, 10676431000119103, 10742121000119104, 708038006, 233678006, 56968009, 2360001000004109, 125011000119100, 195949008, 1086701000000102, 34015007, 10675991000119100, 829976001, 16584951000119101, 782513000, 786836003, 370220003, 735588005, 703953004, 135171000119106, 734905008, 10692681000119108, 1086711000000100, 418395004, 708093000, 10674711000119105, 733858005, 707445000, 401000119107, 708090002, 93432008, 72301000119103, 233683003, 304527002, 424643009, 707981009, 424199006, 10675551000119104, 10674991000119104, 404808000, 407674008, 370218001, 707513007, 404806001, 901000119100, 782520007, 10675391000119101, 703954005, 63088003, 233688007, 1103911000000103, 12428000, 735589002, 1751000119100, 708094006, 389145006, 10675911000119109, 233679003, 16055311000119107, 707512002, 10676391000119108, 707447008, 425969006, 92807009, 10675751000119107, 707979007, 370219009, 18041002, 10692761000119107, 10675871000119106, 708096008, 225057002, 125021000119107, 707444001, 195967001, 866881000000101, 59786004, 442025000, 10692721000119102, 1741000119102, 19849005, 427679007, 707980005, 404804003, 423889005, 31387002, 762521001, 11641008, 370221004, 10675471000119109, 734904007, 55570000, 124991000119109, 708095007, 707511009, 57607007, 281239006, 41553006, 195977004, 409663006, 37981002, 445427006, 427295004, 103781000119103, 10675431000119106, 426979002, 99031000119107, 233687002, 427603009, 10676511000119109, 5281000124103, 135181000119109, 707446004, 125001000119103 |
| inflammatory bowel disease | '295046003', '733157003', '721686000', '128600008', '24829000', '56689002', '71833008', '52506002', '3815005', '50440006', '1085131000119105', '426549001', '737195007', '410485009', '414156000', '61424003', '239814006', '1085901000119101', '235664007', '8161000119106', '444546002', '235714007', '1085231000119100', '201727001', '34000006', '10743231000119101', '697969008', '24526004', '1092841000119100', '1085801000119106', '15342002', '14311001', '414153008', '397173003', '414154002', '201807008', '91390005', '56287005', '239809007', '201805000', '1085851000119105', '196987008', '732966008', '235607002', '13470001', '196977009', '70622003', '1085751000119100', '78324009', '397172008', '444548001', '722850002', '7620006', |

| | |
|---|---|
| | '196578009', '445243001', '441971007', '64766004', '442159003', '234999001', '788718000', '38106008', '78712000', '721702009', '410484008' |
| rheumatoid arthritis | '1073741000119109', '239792003', '11055151000119108', '193180002', '402433007', '1073771000119102', '201772009', '459911000124100', '239804002', '193250002', '735599007', '319031000119108', '1073681000119109', '1073781000119104', '201766009', '201784006', '201780002', '1073791000119101', '410798004', '319841000119107', '201774005', '287008006', '239803008', '201785007', '201768005', '410802003', '201771002', '287006005', '410801005', '38877003', '201778008', '143441000119108', '59165007', '28880005', '15691761000119107', '201783000', '427770001', '399923009', '1073691000119107', '1073811000119102', '15686321000119106', '318941000119109', '239791005', '1073831000119107', '15687841000119108', '86219005', '239795001', '201769002', '201773004', '410796000', '318951000119106', '239799007', '735600005', '15686001000119104', '52661003', '201770001', '15687201000119107', '318871000119107', '410800006', '15685921000119102', '410502007', '15685961000119107', '201779000', '1073711000119105', '201767000', '201777003', '7607008', '319081000119109', '781206002', '77522006', '201775006', '123803006', '62131000000107', '201791009', '15686281000119101', '1073761000119108', '201764007', '1073721000119103', '16024431000119108', '15691161000119108', '410797009', '287007001', '16044751000119106', '239802003', '239941000', '1073731000119100', '57160007', '429192004', '15691801000119104', '15687321000119109', '201776007', '1073821000119109', '15691721000119102', '1073701000119107', '1073751000119106', '300961000119108', '301051000119101', '308143008', '1073801000119100', '398640008', '400054000', '318881000119105', '459921000124108', '410799007', '69896004', '201781003', '319111000119104', '129563009', '201796004' |
| multiple sclerosis | '703621006', '230372003', '439567002', '724778008', '816984002', '426373005', '425500002', '445967004', '24700007', '903741000000102', '733028000', '434321000124107', '428700003', '192929006', '766246000', '192927008', '438511000', '192926004', '703622004' |
| psoriatic arthritis | '239803008', '10629311000119107', '200956002', '239813000', '239802003', '239804002', '410482007', '156370009', '239812005' |
| psoriasis | '238600001', '200969003', '402316001', '402314003', '238616004', '402319008', '402317005', '402326008', '402311006', '402333008', '238611009', '200966005', '402308005', '402336000', '402331005', '238605006', '402330006', '784327005', '238608008', '702617007', '200965009', '200962007', '3533007', '27520001', '200967001', '111188005', '402309002', '402324006', '402318000', '238601002', '402307000', '200968006', '402320002', '28840001', '81271001', '238617008', '238603004', '200975007', '200973000', '238602009', '200972005', '238609000', '200970002', '402312004', '37042000', '238612002', '200971003', '200977004', '239813000', '402315002', '200974006', '238606007', '83839005', '402313009', '402334002', '9014002', '721538000', '33339001', '200963002', '402322005', '402332003', '402323000', '402327004', '200964008', '402325007', '19514005', '238613007', '784339002', '402335001', '402310007', '402321003', '402329001', '719810000', '65539006', '239098009', '238604005', '784328000', '400069004', '402337009', '25847004', '238615000' |
| systemic sclerosis | '196133001', '322461000119108', '236502006', '403519001', '298285004', '236503001', '443872005', '403517004', '402712002', '724603009', '193252005', '774080007', '403520007', '403516008', '715401008', '444133002', '201443009', '22784002', '128461001', '403518009', '87442008', '403514006', '402713007', '35719004', '62382002', '51156002', '89681000119101', '89155008', '299276009', '403515007', '128460000', '31848007' |
| spondyloarthritis | '239806000', '67224007', '239808004', '724606001', '239810002', '235066005', '200956002', '236744002', '234524009', '9350004', '402339007', '162930007', '201776007', '870281008', '1073391000119100', '786077003', '239805001', '328211000119107', '201568008', '723116002', '239813000', '55146009', '1073381000119103', '264516005', '15630971000119102', '10317009', '239815007', '784332006', '713777005', '239811003', '710813008', '15972981000119101', '1074981000119100', '201736002', '9631008', '49807009', '402338004', '239812005', '431236003', '1074971000119103', '371104006', '110041000119104', '15690361000119104', '238407005', '156370009' |
| systemic lupus erythematosus | '25380002', '54912002', '773333003', '309762007', '724767000', '19682006', '402711009', '724781003', '239888002', '397856003', '200936003', '61458006', '403487009', '80258006', '403486000', '77753005', '238652000', '201436003', '698694005', '4676006', '239886003', '196138005', '68815009', '55464009', '54072008', '239889005', '713225000', '239890001', '76521009', '36402006', '200940007', '239887007', '11013005', '73286009', '295121000119101', '22888007', '95609003', '52042003', '707301001', '295101000119105', '95644001', '403488004', '295111000119108' |
| uveitis | '418839003', '818950005', '352941000119102', '416666007', '786077003', '726078000', '699861000', '312930008', '415359008', '410481000', '86219005' |
| vasculitis | '988111000000106', '239926000', '724599009', '359789008', '724502006', '239929007', '128971000119101', '239936008', '402662002', '53485006', '239930002', '232460001', '38675009', '11352009', '58144006', '722020006', '239946005', '28807005', '190815001', '30911005', '57484009', '724783000', '75053002', '191306005', '3275009', '722191003', '310701003', '21542005', '718217000', '195353004', '724600007', '239935007', '239947001', '232369001', '155441006', |

| | |
|---|---|
| | '239927009', '54034003', '235000001', '44371002', '239922003', '403443000', '57390009', '239938009', '723674005', '402856005', '82275008', '239939001', '239945009', '762302008', '230732009', '239925001', '239921005', '239928004' |
| sarcoidosis | '724780002', '402369000', '58870009', '735433009', '697921005', '75403004', '64757003', '233743002', '233744008', '238676008', '402371000', '238680003', '91259005', '402370004', '55941000', '234528007', '54515008', '234524009', '233771008', '707238003', '197368002', '193195000', '232368009', '402372007', '238678009', '111937006', '233769008', '111292008', '195033009', '402380000', '238679001', '9529007', '24369008', '193251003', '37061001', '192673008', '402368008', '233770009', '361197009', '402373002', '231799005', '234526006', '233767005', '72470008', '230193008', '361198004', '400127001', '31541009', '232458003', '233768000', '21787007', '238677004', '234529004', '111936002', '233772001', '234530009', '870334009', '4416007', '234531008', '425384007', '310607007', '80941006', '238674006', '17363001', '234527002', '187233002', '238681004', '402379003', '17919002' |
| antiphospholipid syndrome | '239895006', '239894005', '774084003', '239897003', '239892009', '72161000119100', '402865003', '19267009', '26843008', '609329007' |
| Sjögren's syndrome | '196137000', '239915006', '762303003', '126766000', '83901003', '78946008', '724782005', '239912009' |

**Supplementary Table S9. Specific medications of each immunomodulatory drug class and monoclonal antibody class in this paper.**

The list of actual medications used to define a certain class of immunomodulatory drug in the research paper. Only included the following routes of those medications: 'Oral', 'Intramuscular', 'Intravenous', 'Subcutaneous Infusion', 'Subcutaneous', 'Intravenous (Continuous Infusion)', 'Rectal'.

| Drug class | Example of medications and ingredients | Ingredients RxNorm codes |
|---|---|---|
| hydroxychloroquine | hydroxychloroquine | 5521 |
| methotrexate | methotrexate | 6851 |
| Leflunomide, teriflunomide | leflunomide, teriflunomide | 27169, 1310520 |
| 5-ASA | balsalazide, sulfaSALAzine, mesalamine | 18747, 9524, 52582 |
| azathioprine | azathioprine | 1256 |
| mercaptopurine | mercaptopurine | 103 |
| mitoxantrone | mitoxantrone | 7005 |
| mycophenolate | mycophenolate | 265323 |
| calcineurin inhibitor | cycloSPORINE, sirolimus, tacrolimus | 3008, 35302, 42316 |
| TNF-α inhibitor | adalimumab, certolizumab pegol, etanercept, golimumab, inFLIXimab | 327361, 709271, 214555, 819300, 191831 |
| fumarates | dimethyl fumarate, diroximel fumarate, monomethyl fumarate | 1373478, 2261783, 1546433 |
| interferons | interferon beta-1a, interferon beta-1b | 75917, 72257 |
| alkylating agent | chlorambucil | 2346 |
| hydroxyurea | hydroxyurea | 5552 |
| dapsone | dapsone | 3108 |
| cladribine | cladribine | 44157 |
| IL-1 inhibitor | canakinumab, anakinra, rilonacept | 853491, 72435, 763450 |
| IL-6 inhibitor | sarilumab, tocilizumab, satralizumab | 1923319, 612865, 2391541 |
| IL-12/23 inhibitor | ustekinumab | 847083 |
| IL-17 inhibitor | ixekizumab, brodalumab, secukinumab | 1745099, 1872251, 1599788 |
| IL-23 inhibitor | guselkumab, tildrakizumab, risankizumab | 1928588, 2053436, 2166040 |
| abatacept | abatacept | 614391 |
| anti-BLyS | belimumab | 1092437 |
| S1P receptor modulator | siponimod, ponesimod, fingolimod, ozanimod | 2121085, 2532300, 1012892, 2288236 |
| JAK inhibitor | tofacitinib, upadacitinib, baricitinib | 1357536, 2196092, 2047232 |
| Integrin inhibitor | vedolizumab, natalizumab | 1538097, 354770 |

| | | |
|---|---|---|
| PDE4 inhibitor | apremilast | 1492727 |
| anti-CD20 | riTUXimab, ocrelizumab, ofatumumab | 121191, 1876366, 712566 |
| anti-CD52 | alemtuzumab | 117055 |
| budesonide | budesonide | 19831 |
| systemic glucocorticoids | predniSONE, dexamethasone, prednisoLONE, triamcinolone, methylPREDNISolone, hydrocortisone | 8638, 3264, 8638, 10759, 6902, 5492 |
| Anti-SARS-CoV-2 monoclonal antibody | bamlanivimab, bebelove mab, casirivimab, cilgavimab, etesevimab, etesevimab, imdevimab, sotrovimab, tixagevimab | 2463114, 2592360, 2465242, 2587306, 2477854, 2465249, 2550731, 2587300 |

**Supplementary Table S10. TRIPOD Statement.**

A checklist of items in accordance with TRIPOD reporting guidelines.[1] This includes the page numbers in the manuscript in which the information that fulfils each item on the checklist can be found.

| Section/Topic | | | Checklist Item | Page |
|---|---|---|---|---|
| **Title and abstract** | | | | |
| Title | 1 | D;V | Identify the study as developing and/or validating a multivariable prediction model, the target population, and the outcome to be predicted. | 1 |
| Abstract | 2 | D;V | Provide a summary of objectives, study design, setting, participants, sample size, predictors, outcome, statistical analysis, results, and conclusions. | 1 |
| **Introduction** | | | | |
| Background and objectives | 3a | D;V | Explain the medical context (including whether diagnostic or prognostic) and rationale for developing or validating the multivariable prediction model, including references to existing models. | 2 |
| | 3b | D;V | Specify the objectives, including whether the study describes the development or validation of the model or both. | 3-10 |
| **Methods** | | | | |
| Source of data | 4a | D;V | Describe the study design or source of data (e.g., randomized trial, cohort, or registry data), separately for the development and validation data sets, if applicable. | 3-5; Figure 1 |
| | 4b | D;V | Specify the key study dates, including start of accrual; end of accrual; and, if applicable, end of follow-up. | 3-4; Figure 1-2 |
| Participants | 5a | D;V | Specify key elements of the study setting (e.g., primary care, secondary care, general population) including number and location of centres. | 3-4 |
| | 5b | D;V | Describe eligibility criteria for participants. | 3; Figure 1 |
| | 5c | D;V | Give details of treatments received, if relevant. | 5-9; Table 2 |
| Outcome | 6a | D;V | Clearly define the outcome that is predicted by the prediction model, including how and when assessed. | 3 |
| | 6b | D;V | Report any actions to blind assessment of the outcome to be predicted. | 3-5 |

| | | | | |
|---|---|---|---|---|
| Predictors | 7a | D;V | Clearly define all predictors used in developing or validating the multivariable prediction model, including how and when they were measured. | 5-9; Table 2 |
| | 7b | D;V | Report any actions to blind assessment of predictors for the outcome and other predictors. | 3-5 |
| Sample size | 8 | D;V | Explain how the study size was arrived at. | 3-4; Figure 1 |
| Missing data | 9 | D;V | Describe how missing data were handled (e.g., complete-case analysis, single imputation, multiple imputation) with details of any imputation method. | 4; Table 2 |
| Statistical analysis methods | 10a | D | Describe how predictors were handled in the analyses. | 3-4 |
| | 10b | D | Specify type of model, all model-building procedures (including any predictor selection), and method for internal validation. | 3-5 |
| | 10c | V | For validation, describe how the predictions were calculated. | 4; Figure 2 |
| | 10d | D;V | Specify all measures used to assess model performance and, if relevant, to compare multiple models. | 3-5; Table S1 |
| | 10e | V | Describe any model updating (e.g., recalibration) arising from the validation, if done. | 3-5; Table S1 |
| Risk groups | 11 | D;V | Provide details on how risk groups were created, if done. | NA |
| Development vs. validation | 12 | V | For validation, identify any differences from the development data in setting, eligibility criteria, outcome, and predictors. | 3-4 |
| **Results** | | | | |
| Participants | 13a | D;V | Describe the flow of participants through the study, including the number of participants with and without the outcome and, if applicable, a summary of the follow-up time. A diagram may be helpful. | 3-4; Figure 1 |
| | 13b | D;V | Describe the characteristics of the participants (basic demographics, clinical features, available predictors), including the number of participants with missing data for predictors and outcome. | 5-9; Table 2 |
| | 13c | V | For validation, show a comparison with the development data of the distribution of important variables (demographics, predictors and outcome). | 5-9; Table 1-2 |
| Model development | 14a | D | Specify the number of participants and outcome events in each analysis. | 3-5; Table 1 |
| | 14b | D | If done, report the unadjusted association between each candidate predictor and outcome. | N/A |
| Model specification | 15a | D | Present the full prediction model to allow predictions for individuals (i.e., all regression coefficients, and model intercept or baseline survival at a given time point). | N/A |
| | 15b | D | Explain how to the use the prediction model. | N/A |
| Model performance | 16 | D;V | Report performance measures (with CIs) for the prediction model. | 6; Table S3-S8 |

| | | | | |
|---|---|---|---|---|
| Model-updating | 17 | V | If done, report the results from any model updating (i.e., model specification, model performance). | Figure S1 |
| **Discussion** | | | | |
| Limitations | 18 | D;V | Discuss any limitations of the study (such as nonrepresentative sample, few events per predictor, missing data). | 11 |
| Interpretation | 19a | V | For validation, discuss the results with reference to performance in the development data, and any other validation data. | 7-11 |
| | 19b | D;V | Give an overall interpretation of the results, considering objectives, limitations, results from similar studies, and other relevant evidence. | 7-11 |
| Implications | 20 | D;V | Discuss the potential clinical use of the model and implications for future research. | 11 |
| **Other information** | | | | |
| Supplementary information | 21 | D;V | Provide information about the availability of supplementary resources, such as study protocol, Web calculator, and data sets. | Y |
| Funding | 22 | D;V | Give the source of funding and the role of the funders for the present study. | 12 |

**Supplementary Table S11. STROBE Statement.**

A checklist of items in accordance with STROBE reporting guidelines.[2] This includes the page numbers in the manuscript in which the information that fulfils each item on the checklist can be found.

| | Item No· | Recommendation | Page No· | Relevant text from manuscript |
|---|---|---|---|---|
| **Title and abstract** | 1 | (*a*) Indicate the study’s design with a commonly used term in the title or the abstract | 1 | Risk factors for severe COVID-19 outcomes: a study of immune-mediated inflammatory diseases, immunomodulatory medications, and comorbidities in a large US healthcare system; Title |
| | | (*b*) Provide in the abstract an informative and balanced summary of what was done and what was found | 1 | Abstract |
| **Introduction** | | | | |
| Background/rationale | 2 | Explain the scientific background and rationale for the investigation being reported | 2 | Introduction |

| | | | | |
|---|---|---|---|---|
| Objectives | 3 | State specific objectives, including any prespecified hypotheses | 2 | Here, we analyse the risk for severe COVID-19 outcomes, using multivariable models across a large US population to investigate how specific IMIDs, classes of immunomodulatory drugs, chronic comorbidities, and vaccination status are associated with risk for severe COVID-19 outcomes, and compare the pre-Omicron and Omicron-predominant time periods· |
| **Methods** | | | | |
| Study design | 4 | Present key elements of study design early in the paper | 2 | Using multivariable models across a large US population |
| Setting | 5 | Describe the setting, locations, and relevant dates, including periods of recruitment, exposure, follow-up, and data collection | 3-6 | Method |
| Participants | 6 | (*a*) *Cohort study*—Give the eligibility criteria, and the sources and methods of selection of participants· Describe methods of follow-up<br>*Case-control study*—Give the eligibility criteria, and the sources and methods of case ascertainment and control selection· Give the rationale for the choice of cases and controls<br>*Cross-sectional study*—Give the eligibility criteria, and the sources and methods of selection of participants | 3 and 4 | *Study setting and participants; Cohort characteristics* |
| | | (*b*) *Cohort study*—For matched studies, give matching criteria and number of exposed and unexposed<br>*Case-control study*—For matched studies, give matching criteria and the number of controls per case | N/A | This was an unmatched study |
| Variables | 7 | Clearly define all outcomes, exposures, predictors, potential confounders, and effect modifiers· Give diagnostic criteria, if applicable | 3 and 4 | *COVID-19 severe outcomes;* Figure 1 |
| Data sources/ measurement | 8* | For each variable of interest, give sources of data and details of methods of assessment (measurement)· Describe comparability of assessment methods if there is more than one group | 3 and 4 | Figure 1; Supplementary Table S9; Supplementary Table S10 |

| | | | | |
|---|---|---|---|---|
| Bias | 9 | Describe any efforts to address potential sources of bias | 3-6 | Models were trained for two SARS-CoV-2 positive cohorts (those infected before the pre-dominance of Omicron variants and those infected later)·Models were trained on 90% of the data with (over-sampling or over-weight on minority classes to address class imbalance in training data), with 10% of the data held out for independent performance testing· |
| Study size | 10 | Explain how the study size was arrived at | 3 and 4 | Figure 1 |
| Quantitative variables | 11 | Explain how quantitative variables were handled in the analyses· If applicable, describe which groupings were chosen and why | 3-5 | *Machine learning models used for classification* |
| Statistical methods | 12 | (*a*) Describe all statistical methods, including those used to control for confounding | 3-6 | *Machine learning models used for classification* |
| | | (*b*) Describe any methods used to examine subgroups and interactions | N/A | - |
| | | (*c*) Explain how missing data were addressed | 4 | *Machine learning models used for classification* |
| | | (*d*) *Cohort study*—If applicable, explain how loss to follow-up was addressed<br>*Case-control study*—If applicable, explain how matching of cases and controls was addressed<br>*Cross-sectional study*—If applicable, describe analytical methods taking account of sampling strategy | 3 | *Study setting and participants* |
| | | (*e*) Describe any sensitivity analyses | 6 | *Machine learning models used for classification* |
| **Results** | | | | |

| | | | | |
|---|---|---|---|---|
| Participants | 13* | (a) Report numbers of individuals at each stage of study—eg numbers potentially eligible, examined for eligibility, confirmed eligible, included in the study, completing follow-up, and analysed | 3 | *Study setting and participants* |
| | | (b) Give reasons for non-participation at each stage | N/A | - |
| | | (c) Consider use of a flow diagram | 4 | Figure 1 |
| Descriptive data | 14* | (a) Give characteristics of study participants (eg demographic, clinical, social) and information on exposures and potential confounders | 6 and 7 | Table 1 and Table 2 |
| | | (b) Indicate number of participants with missing data for each variable of interest | N/A | There was no missing data after imputation |
| | | (c) *Cohort study*—Summarise follow-up time (eg, average and total amount) | N/A | This was a retrospective study |
| Outcome data | 15* | *Cohort study*—Report numbers of outcome events or summary measures over time | 3 | Table 1 |
| | | *Case-control study*—Report numbers in each exposure category, or summary measures of exposure | N/A | This was a retrospective study |
| | | *Cross-sectional study*—Report numbers of outcome events or summary measures | N/A | This was a retrospective study |
| Main results | 16 | (*a*) Give unadjusted estimates and, if applicable, confounder-adjusted estimates and their precision (eg, 95% confidence interval)· Make clear which confounders were adjusted for and why they were included | 6 and 7 | Results |
| | | (*b*) Report category boundaries when continuous variables were categorized | 3 and 4 | Continuous variables were normalized by applying min-max transformation |
| | | © If relevant, consider translating estimates of relative risk into absolute risk for a meaningful time period | N/A | There were no estimates of relative risk |
| Other analyses | 17 | Report other analyses done—eg analyses of subgroups and interactions, and sensitivity analyses | N/A | There were no subgroups |

| **Discussion** | | | | |
|---|---|---|---|---|
| Key results | 18 | Summarise key results with reference to study objectives | 7 | Discussion |
| Limitations | 19 | Discuss limitations of the study, taking into account sources of potential bias or imprecision· Discuss both direction and magnitude of any potential bias | 7 | Discussion |
| Interpretation | 20 | Give a cautious overall interpretation of results considering objectives, limitations, multiplicity of analyses, results from similar studies, and other relevant evidence | 7-11 | Discussion |
| Generalisability | 21 | Discuss the generalisability (external validity) of the study results | 11 | Discussion; In strength and limitation paragraphs |
| **Other information** | | | | |
| Funding | 22 | Give the source of funding and the role of the funders for the present study and, if applicable, for the original study on which the present article is based | 1 | Abstract |

**Supplementary Table S12. Classification Machine learning algorithms hyperparameters and hyperparameter values used in performance evaluation.**

| Model name | Name | Value |
|---|---|---|
| Logistic Regression | missing | (‘drop’) |
| | method | (‘lbfgs’) |
| K Nearest Neighbours | n_neighbors | (10) |
| | weights | (‘uniform’) |
| | algorithm | (‘auto’) |
| | leaf_size | (30) |
| | p | (2) |
| Support Vector Machine | kernel | (‘rbf’) |
| | C | (1) |
| | gamma | (‘scale’) |
| | shrinking | (True) |
| | probability | (True) |
| | class_weight | (‘balanced’) |
| | random_state | (42) |
| Adaptive Boosting | n_estimators | (200) |
| | learning_rate | (0.05) |
| | algorithm | ('SAMME.R’) |
| | random_state | (42) |

### Supplementary Discussion

**Supplementary Discussion S1. Anti-SARS-CoV-2 neutralizing monoclonal antibodies (NmAbs) analysis.**

Anti-SARS-CoV-2 NmAbs were only administered to 0·41% (699 / 169 993) of patients with positive COVID-19 tests during the pre-Omicron period, and only 2·34% (2 822 / 120 862) during the Omicron pre-dominant period. (Table 2). NmAbs were significantly associated with reduced risk for severe outcomes in both periods. Both the NmAbs analysis and the analysis excluding patients who received NmAbs showed similar results to the main analyses for other variables. The additional analyses on NmAbs showed that few patients received this treatment. NmAbs have been recommended for patients who test positive and are considered at high risk but do not yet require hospitalisation. Because IMIDs and immunomodulatory medications have sometimes been assumed to confer high risk, the significant association between NmAbs and better outcomes may reflect a combination of treatment efficacy and confounding by indication.

**Supplementary Discussion S2. Paxlovid analysis.**

Paxlovid was only administered to 0·12% (146 / 120 862) of patients with positive COVID-19 tests during the Omicron pre-dominant period. (Table 2). Paxlovid did not reach significance during the Omicron time period which it is studied.
